# Surveying the Genomic Landscape of Mantle Cell Lymphoma Indicates the Importance of Multimodal Genomic and Transcriptomic Exploration

**DOI:** 10.64898/2026.07.25.26358867

**Authors:** Charlz Nithin Jerold, Brian Li, Matthew Mosior, David Russler-Germain, Anshu Dahal, Zachary Skidmore, Kelsy C. Cotto, Malachi Griffith, Todd A. Fehniger, Obi L. Griffith, Brad Kahl, Felicia Gomez

**Affiliations:** Washington University School of Medicine, Department of Medicine, St. Louis, MO 63108; McDonnell Genome Institute, Department of Medicine, Washington University School of Medicine, St Louis, MO; Department of Genetics, Washington University School of Medicine, St Louis, MO; Siteman Cancer Center, Washington University School of Medicine, St Louis, MO

**Keywords:** Mantle cell lymphoma, exome, RNA-seq, whole genome sequencing

## Abstract

Mantle cell lymphoma (MCL) is a B-cell non-Hodgkin lymphoma characterized by heterogeneous clinical courses despite a common pathobiological initiating event. In this work we explore the genomic variants that characterize MCL and integrate transcriptomic data to comprehensively describe MCL biology. We performed whole exome sequencing (WES) on 28 tumor-normal pairs (lymph node and skin, respectively), as well as whole genome sequencing (WGS) and RNA sequencing on subsets of samples. We used established DNA and RNA analysis pipelines to detect single-nucleotide variants (SNV) and indels, structural variants, copy-number alterations, and RNA fusions. The canonical t(11;14)(q13;q32) *CCND1::IGH* translocation was detected in 8 of 10 WGS samples. Structural variant analysis additionally identified recurrent rearrangements involving *KMT2A* and *PAFAH1B2*. SNV and indel analyses revealed frequent mutations in *ATM, TP53, CCND1, IGH*, and *NOTCH1*. *ATM* exhibited diverse variant classes, including missense mutations, frameshift mutations, deletions, and duplications, while all detected *NOTCH1* mutations were predicted loss-of-function frameshift variants. Copy-number analysis identified recurrent losses affecting DNA damage response genes, including *TP53* and *ATM*, and recurrent gains involving transcriptional regulators and oncogenic signaling genes. Integrated pathway analysis demonstrated enrichment of transcriptional misregulation, DNA repair, *PI3K/AKT* signaling, and interleukin signaling pathways. We also identified recurrent alterations in candidate genes, including *ASXL1*, suggesting additional mechanisms of epigenetic dysregulation in MCL.

Together, these findings provide a comprehensive description of somatic alterations in MCL and demonstrate that diverse genomic lesions converge on common pathways involved in genomic instability, transcriptional regulation, and tumor survival.

## Introduction

Mantle cell lymphoma (MCL) is an aggressive mature B-cell non-Hodgkin lymphoma. It originates when mature B lymphocytes in the mantle zone of a lymph node follicle acquire mutations that lead to aberrant proliferation.^1^ MCL is characterized by the t(11;14)(q13;q32) translocation, in which the gene encoding cyclin D1 (*CCND1*) from 11q13 is aberrantly associated by the *IGH* locus’s enhancer elements at 14q32 ^2,3^ This lesion results in overexpression of cyclin D1, ultimately leading to reduced apoptosis and dysregulation of the cell cycle.^4^ While the vast majority of MCL cases harbor this translocation, the presence of secondary mutations are known to induce further genetic instability and contribute to tumorigenesis. Some of these established recurrently mutated genes in MCL include *ATM*, *CCND1*, *TP53*, *KMT2D*, *NOTCH1*, and *NSD2* ^5,6^ These alterations span multiple variant classes, including single-nucleotide variants (SNVs), small insertions and deletions (indels), structural variants, and large-scale copy number alterations ^7^. Although recent therapeutic advances have substantially improved outcomes for many patients with MCL, clinical outcomes remain heterogeneous and are influenced by the underlying genomic landscape. Patients harboring high-risk genomic features, including TP53 alterations, 17p deletions, and complex karyotypes, continue to experience inferior outcomes compared with other molecular subgroups. This clinical and molecular heterogeneity highlights the need for comprehensive genomic characterization to improve biological understanding, refine risk stratification, and identify additional therapeutic targets.^8^

Frontline treatment of MCL typically consists of chemoimmunotherapy with the anti-CD20 monoclonal antibody rituximab plus with either bendamustine, anthracycline-based, or cytarabine-based combinations. Recent clinical trials have explored combining these backbones with targeted therapies intended to inhibit specific MCL dependencies including B-cell receptor signaling, the PI3K/AKT/mTOR pathway, checkpoints in the DNA repair pathway, anti-apoptotic pathways, and the ubiquitin-proteasome network, among others.^9^ Despite these advances, patients with high-risk genomic features, particularly *TP53* alterations, continue to experience inferior outcomes with conventional chemoimmunotherapy. Consequently, emerging chemotherapy-free approaches, such as zanubrutinib, obinutuzumab, and venetoclax (BOVen), have demonstrated promising efficacy in TP53-mutated MCL, highlighting the growing importance of genomic profiling in guiding therapeutic selection. ^10^

In the current study, we explore a variety of genomic mutation types and transcriptomic variation in MCL patients through the use of whole exome sequencing (WES), whole genome sequencing (WGS), and RNA sequencing (RNA-seq). We report novel somatic mutations (indels, single nucleotide variants (SNVs) and structural variants) and describe RNA fusion events in MCL. Our study demonstrates the promise of conducting mutational analyses across different sequencing technologies to better characterize the comprehensive set of genetic mutations in MCL patients.

## Methods

### Patient Samples

The present study analyzed fresh frozen tumors (lymph nodes or bone marrow) from 28 patients with MCL (Table1). Non-malignant samples were collected (skin punch biopsies) and were included for germline analysis. Frozen sections (tumor and skin) were cut and used for genomic DNA isolation. All patients provided written informed consent for the use of their samples in sequencing as part of the Washington University School of Medicine (WUSM) Lymphoma Banking Program. The WUSM Institutional Review Board (IRB) approved protocols include: IRB 201108251, 201104048, 201110187. All human research activities are guided by the ethical principles in “The Belmont Report: Ethical Principles and Guidelines for the Protection of Human Subjects Research of the National Commission for the Protection of Human Subjects of Biomedical and Behavioral Research”.

### DNA and RNA Isolation

DNA and RNA was isolated at the Washington University Tissue Procurement Core facility using standard procedures (https://sites.wustl.edu/sitemanbbr/).

### SNV/Indel calling, filtering, and manual review

Sequence analysis and data management was performed using the Genome Modeling System (GMS)^11^. Briefly, paired-end reads were aligned to the human reference sequence GRCh38 using BWA-MEM v0.7.10-r789^12^. Duplicate marking was completed using samblaster v0.1.22^13^ and alignments were sorted using sambamba v0.5.4^14^. Somatic variants were called using SAMtools^15^, SomaticSniper^16^, VarScan^17^, MuTect^18^, Strelka^19^, Pindel^20^, and GATK^21^. Variants were required to be called by at least one variant caller. After variant calling, all variants underwent filtering and manual review. The filtering pipeline consisted of filters based on read depth and the number of variant supporting reads (≥20 read depth in tumor/normal, ≥5 variant supporting reads, <5% normal VAF), Ensembl consequence filtering, and gnomAD subpopulation filtering (<1% maximum allele frequency in all gnomad subpopulations) The remaining variants were manually reviewed in IGV^22^.

### SNV/Indel Analysis

#### Co-Occurence and Mutual Exclusivity

When we tested for significant interactions between SNVs and indels using Fisher’s exact test. Associations with *p* < 0.05 were considered statistically significant; results with *p* < 0.01 were noted as stronger evidence of association.

#### Significant mutated genes analysis

To identify significantly mutated genes, in our MCL cohort, we applied the dN/dScv R package (v0.0.1.0). The input consisted of somatic SNVs identified through our whole-exome sequencing (WES) analysis. Each variant record included the chromosome, position, reference, and alternate allele, and was restricted to coding substitutions annotated as either synonymous (to establish a background mutation rate) or nonsynonymous.

Analysis was performed using the hg38 reference genome, and model fitting was conducted using maximum-likelihood estimation to compute normalized dN/dS (ω) ratios. Global and gene-level estimates were derived from the dndscv() output, including globaldnds, sel_cv, and annotmuts tables. Genes were considered to show a rate of mutation that is elevated with respect to the background mutation rate (evidence of positive selection) if their qglobal_cv < 0.05 and recurrent in 2 samples. This framework provides a quantitative measure of increased mutation rate in the context of identifying selective pressure. The goal is to identify genes in which nonsynonymous substitutions occur at higher-than-expected rates relative to synonymous substitutions, indicative of potential oncogenic driver activity.

#### Copy Number Variant Calling

Copy number variants were identified using CNVkit. This software was applied to the 27 WES samples and 10 WGS samples. CNVkit segmentation was run using a 100kb target bin size on both the WGS and WES data. Copy-number segments were filtered based on size (>10 Mb) and log-scale copy-number amplitude (>0.4 or <–0.4), corresponding to absolute copy numbers >3 or <1. This approach retained only large chromosomal regions exhibiting clear copy-number gains or losses. The resulting copy number segments were filtered based on size (> 10Mb) and copy number level in log scale (> 0.4 or < -0.4), which means that the absolute copy number was > 3 or < 1 resulting in large-scale chromosomal regions that were significantly copy-number altered meaning showed either gain or loss. The copy-number altered regions were aggregated between the WES and WGS samples. Additionally, as a simplifying procedure we estimated the recurrence based on mutated chromosome arms. While the WES and WGS copy number analyses were separate and had an unequal number of samples, CN events in the WGS samples provide further resolution and clarify, validate, or refine the corresponding CN events detected in the WES samples

#### Structural Variant Calling

Structural variant analysis was conducted using the Manta^23^ on the 10 WGS samples. The SVs called by Manta were first filtered based on the number of reads (≥10 total tumor supporting reads) and then manually reviewed using svviz^24^ and IGV. To estimate the impact of an SV 500kb flank was added to the SVs not removed by filtering.

Analysis of genes affected by structural variants was restricted to genes that were previously implicated in cancer, according to the COSMIC Cancer Gene Census.

#### RNA fusion Calling

RNA fusion analysis was conducted using STAR-Fusion on the 8 RNA-Seq samples. Fusion transcripts were filtered based on the total number of junction and spanning reads that supported the fusion (≥10 total junction and spanning reads). Following this filtering step all fusions were manually reviewed using FusionInspector.^25^ Fusions involving genes that are right beside each other were labeled as readthrough transcripts. These occur when transcription continues past the normal end of one gene into the next, creating an apparent fusion without any underlying genomic rearrangement. Because they do not represent true fusion events, they were excluded from further analysis.

#### Pathway Enrichment Analysis

To evaluate the biological significance of the identified somatic alterations, we performed pathway enrichment analysis using the aggregated set of mutated genes from all variant classes, including single-nucleotide variants (SNVs), small insertions/deletions (indels), structural variants, and copy number alterations. Functional enrichment testing was performed against curated KEGG and Reactome pathway databases. Gene symbols were mapped to Entrez Gene identifiers using *org.Hs.eg.db* to ensure standardized annotation across tools. Enrichment was assessed using a two-sided Fisher’s exact test as implemented in R packages such as *clusterProfiler (v4.14.0)*^26^ and *gage (v2.56.0)*^27^, comparing the frequency of altered genes within each pathway against the genome-wide background of non-mutated genes. Multiple-testing correction was applied using the Benjamini–Hochberg false discovery rate (FDR) procedure, with FDR < 0.05 considered statistically significant. Significant pathways were further categorized according to the most number of mutations present in each (copy-number gain, copy-number loss, or point mutation). Pathway visualizations and gene–pathway overlays were generated using R packages such as *pathview(v1.46.0)*^28^ and *ReactomePA(1.50.0)*^29^ to highlight genes recurrently perturbed across the cohort.

## Results

### Sequencing Overview and Quality Control

All 28 MCL patient samples underwent multi-platform sequencing, including 28 tumor–normal pairs analyzed by WES, 10 samples subjected to WGS- tumor only, and 8 samples profiled using RNA-seq. The WES data achieved an average per-base coverage of 111.7x for normal samples and 124.4x for tumor samples. The WGS data achieved 31.1x and 26.0x mean coverage for normal and tumor samples, respectively. The RNA-seq data demonstrated high quality, with an average of 96.9% of reads mapping to transcriptome and a mean of 2.07 × 10⁸ total reads per tumor sample.

Quality control of the WES models using the Somalier relatedness tool identified one patient (TWGE-08-0075-1027) with unexpectedly low genetic concordance between the tumor and matched normal sample (Supplemental Figure 1). This case was excluded from subsequent analyses due to suspected cross-sample contamination. ^30^

These sequencing metrics confirm that the majority of samples met high-coverage and quality thresholds necessary for accurate detection of single-nucleotide variants, indels, structural variants, and copy-number alterations.

### Recurrent SNVs and indels

Somatic mutation analysis was performed on 27 WES samples (see Table 1). Initial variant calls underwent filtering and manual review to create a final set of high quality somatic mutations (see Methods). In our patient cohort, we identified 588 SNVs and indels. Missense variants were the most common type of variant (494/588), followed by stop gained variants (40/588) and frameshift variants (39/588; Supplemental Table 1 and Supplemental Figure 2). Overall, this mutational burden is modest and consistent with what has been reported for MCL, which typically exhibits a lower mutation rate compared to more genomically complex lymphomas such as diffuse large B-cell lymphoma (DLBCL) and, to a lesser extent, follicular lymphoma (FL). Each patient had an average of 21 SNVs and indels (range: 0–61) with a mean variant allele frequency (VAF) of approximately 27.5% (median, 28.0%; range, 2.5–92.4%).Sample MCL 930 was the only sample with 0 identified SNVs and indels that passed filtering and manual review, but it was found to still harbor other types of mutations, such as copy number losses in chromosome 13 region 13q31.3 to 13q34 affecting *GPC5*, *SOX21*, and *ERCC5*, and in chromosome 17 region 17p13.3 to 17p11.2 affecting *USP6*, *RABEP1*, *TP53*, *PER1*, *GAS7*, *MAP2K4* and *NCOR1*. SNVs and indels present in at least two samples are summarized in Supplemental Figure 3. These variants impacted a total of 25 genes, 11 of which have been previously reported as mutated in MCL, including *ATM* (7/27; 26%), *TP53* (6/27; 22%), *CCND1* (4/27; 15%), *NSD2* (3/27; 11%), and *NOTCH1* (3/27; 11%). In addition to these recurrently mutated genes, we observed non-recurrent mutations in several genes with established relevance to B-cell receptor and NF-κB signaling, including *CD79B, CARD11, and PLCG2,* as well as chromatin-regulatory genes including *SMARCA4*. Although these alterations occurred below the recurrence threshold used for Supplemental Figure 3, their known biological roles are relevant to MCL biology and are therefore considered in the pathway-level interpretation below. We identified recurrent somatic SNVs or indels in 14 genes not previously reported as recurrently mutated in MCL cohorts, each occurring in two cases (2/27; 7.4%), including *ADGRE1*, *ANKRD36C*, *ASXL1*, *FAM120C*, *GABRD*, *HRNR*, *KIAA1551*, *MPP1*, *MUC12*, *PCDH15*, *SEMA5A*, *TNC*, *WDFY3*, and *ZNF804B*. The role of *ASXL1* variants in MCL is uncharacterized. We identified two potential loss-of-function mutations in exon 13 (Supplemental Figure 4). *ASXL1* has been found to be involved in other cancers, including chronic myeloid leukemia.^31^ ^32^ *ZNF804B* is also a novel mutated gene identified here, with three novel missense variants across two samples.

**Table 1:** Patient characteristics of the 27 samples in the MCL cohort. Data are presented as number (%) unless otherwise indicated. Frontline therapies include bendamustine plus rituximab (BR), rituximab plus methotrexate with cyclophosphamide, doxorubicin, vincristine, and prednisone (RM-CHOP), and rituximab plus hyper-cyclophosphamide, vincristine, Adriamycin, and dexamethasone (R-Hyper-CVAD).

| Characteristic |  |
| --- | --- |
| Total Patient Number | 27 |
| Male (%) | 20 (74.1) |
| Age (Mean) | 63 |
| Age range | 36-84 |
| **Stage (%)** |  |
| I | 0 (0) |
| II | 1 (3.7) |
| III | 2 (7.4) |
| IV | 24 (88.9) |
| **ECOG Status (%)** |  |
| 0 | 20 (74.1) |
| 1 | 5 (18.5) |
| 2 | 1 (3.7) |
| 3 | 1 (3.7) |
| **Treatment Stage at Biopsy (%)** |  |
| Pre-treatment | 19 (70.4) |
| 1st Relapse | 4 (14.8) |
| 2nd Relapse | 2 (7.4) |
| 3rd Relapse | 2 (7.4) |
| **Frontline Treatment (%)** |  |
| BR | 15 (55.6) |
| RM-CHOP | 5 (18.5) |
| Observation | 5 (18.5) |
| R-Hyper-CVAD | 2 (7.4) |

Blastoid or pleomorphic morphology was documented in 5 of 27 patients. Alterations in previously reported high-risk MCL-associated genes were observed in 4 of these 5 cases, most commonly involving *TP53* and *NOTCH1*. Given the small number of blastoid/pleomorphic cases, this observation is descriptive and is not sufficient to establish a statistically significant association.^33^

Interactions between recurrent SNVs and indels were analyzed to identify co-occurring or mutually exclusive mutation pairs. Eleven gene pairs showed significant co-occurrence (p < 0.05; Supplemental Figure 5), including the cell cycle regulators *CCND1* and *TP53*, which were co-mutated in 3 patients (p < 0.05). No mutually exclusive pairs reached statistical significance. Because *ATM* and *TP53* were also affected by copy-number alterations, we summarized their alteration patterns across both SNV/indel and copy-number events. When both alteration classes were considered, *ATM* was altered in 8 of 27 samples and *TP53* was altered in 9 of 27 samples. Six samples only had *ATM* alterations, seven only had *TP53* alterations, and two samples harbored alterations affecting both genes. These findings indicate limited overlap between *ATM* and *TP53* alterations in this cohort, but do not support statistically significant mutual exclusivity.

Previous work ^34^ showed that splice site mutations in *HNRNPH1* were mutated in 10% of MCL patients and impact alternative splicing of *HNRNPH1* transcripts. These prior studies also showed that these alternative transcripts result in increased expression of HNRNPH1 through reduced autoregulation ^34^. We searched for these mutations in our patients, finding *HNRNPH1* splice site mutations in 2 of 27 WES samples (> 0.05 VAF and variant support read count > 5), both of which had been previously reported. ^34^ The coverage for all *HNRNPH1* splice-site mutations interrogated is summarized in Supplemental Table 2. Overall, the coverage at each *HNRNPH1* site that we interrogated is consistent with the average reported coverage for the exome and whole genome data. Exome sequencing depth across the interrogated splice-site loci was consistently high (typically >130×; Supplemental Table 2), indicating that the low frequency of *HNRNPH1* splice-site mutations in our cohort is unlikely to reflect insufficient sequencing coverage.

### Significant mutated genes analysis

Somatic coding mutations were analyzed using the dN/dS framework implemented in dndscv to identify genes under positive selection, defined by an excess of non-synonymous relative to synonymous mutations. Across the cohort, *TP53* was the only gene showing statistically significant evidence of positive selection (q < 0.05), whereas *ATM* demonstrated a borderline signal that did not reach significance (wmis = 48.919, q = 0.06), likely reflecting the limited statistical power of the cohort.

Independent of the dN/dS analysis, the most frequently mutated genes were *ATM* (8/27, 29.6%) and *TP53* (7/27, 25.9%), followed by *CELA1* (3/27, 11.1%), *DAZAP1* (3/27, 11.1%), and *ASXL1* (2/27, 7.4%) (Supplemental Table 3). Although recurrently mutated, these latter genes (*CELA1*, *DAZAP1*, and *ASXL1*) did not show statistically significant evidence of positive selection in the dndscv analysis. (Supplemental Table 3)

### Recurrent Structural Variants and Copy Number Events

Structural variant analysis on the 10 WGS samples revealed 142 total events, consisting of 76 translocation, 48 deletion, and 18 tandem duplications. The most recurrent structural variant was the canonical t(11;14)(q13;q32) chromosomal translocation (Figure 1), which was detected in 8 of 10 samples. No structural rearrangements involving *CCND2* or *CCND3* were identified in the remaining WGS samples. Other recurrent structural events included intrachromosomal translocations on chromosome 2 (3/10; 30%) involving the *IGK* locus and deletions on chromosome 11 (2/10; 20%) affecting *KMT2A* and *PAFAH1B2*. (Supplemental Table 4 and Supplemental Figure 6).

**Figure 1:**
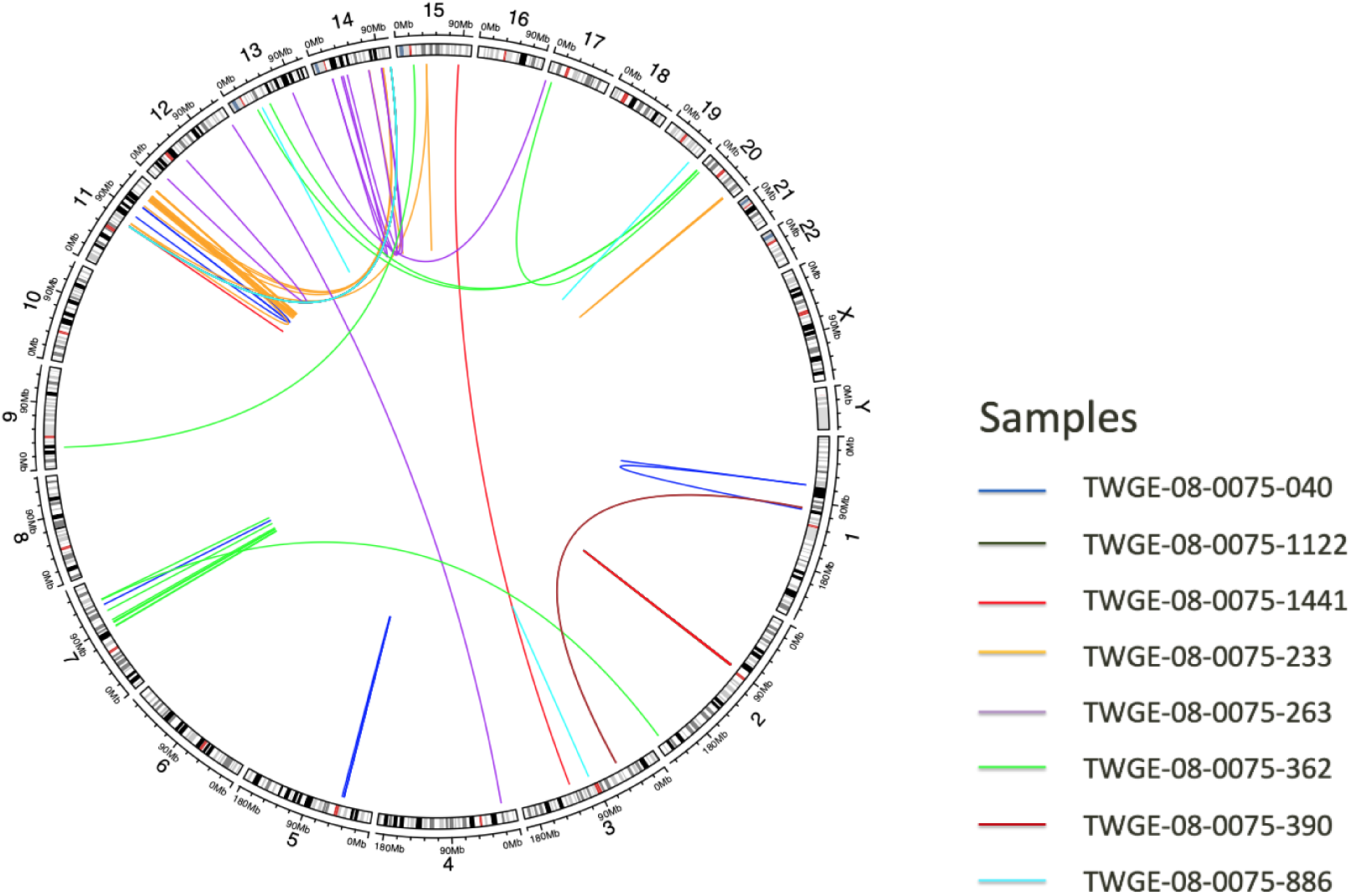
Translocation Circos Plot. Translocations detected in samples for which whole genome data was available (n=10). 8 of the WGS samples harbored at least one chromosomal translocation. Translocations are depicted using a line connecting the adjoining chromosomal locations. Each color represents a unique sample.

When we combined copy number analysis using WES and WGS data, we found 37 unique events where a chromosomal arm was gained or lost in at least one sample. The most recurrent copy-number gained chromosomal arms were 3q (5/27; 18.5%), 7p (4/27; 14.8%), 15q (4/27; 14.8%), and 8q (2/27; 7.4%). The most recurrent copy-number deleted chromosomal arms were 13q, 15q, 9q, 11q, 17p, 6q, and 8p (Table 2). All of the most common copy-number events (n >= 2) impacted genes known to be mutated in MCL including *ATM*, *TP53*, and *RB1*^35,36^ (Supplemental Table 5)

**Table 2:** Recurrent copy-number events summarized at the chromosome arm level (n=27). This table summarizes copy number events identified in the exome data as well as whole genome data, when available. Chromosome arms with at least two mutated samples were included in the table.

| Chromosome Arm | CN Event | Affected Genes | Patients | Frequency |
| --- | --- | --- | --- | --- |
| 13q | Loss | ZMYM2,<br>LATS2, CDX2,<br>FLT3, BRCA2,<br>NBEA,<br>LHFPL6,<br>FOXO1, LCP1,<br>RB1,<br>CYSLTR2,<br>GPC5, SOX21,<br>ERCC5 | 6 | 0.22 |
| 3q | Gain | TFG, CBLB,<br>POLQ, GATA2,<br>RPN1, CNBP,<br>STAG1,<br>PIK3CB,<br>FOXL2, ATR,<br>WWTR1,<br>GMPS, MLF1,<br>MECOM,<br>TBL1XR1,<br>PIK3CA,<br>SOX2,<br>MAP3K13,<br>IGF2BP2,<br>ETV5, EIF4A2,<br>BCL6, LPP,<br>TP63,<br>MB21D2,<br>MUC4, TFRC | 5 | 0.19 |
| 15q | Loss | NUTM1,<br>BUB1B,<br>KNSTRN,<br>NTRK3,<br>POLG, IDH2,<br>CRTC3, BLM,<br>FES, CHD2,<br>KNL1 | 4 | 0.15 |
| 7p | Gain | ETV1, MACC1,<br>HNRNPA2B1,<br>HOXA9,<br>HOXA11,<br>HOXA13,<br>JAZF1,<br>FKBP9,<br>SFRP4, IKZF1,<br>EGFR,<br>ZNF479,<br>CARD11,<br>PMS2, RAC1 | 4 | 0.15 |
| 9q | Loss | GNAQ, SYK,<br>ABL1,<br>NUP214,<br>TSC1,<br>RALGDS,<br>BRD3,<br>NOTCH1,<br>FNBP1 | 4 | 0.15 |
| 11q | Loss | FAT3, MANBA,<br>BIRC3, ATM,<br>DDX10,<br>POU2AF1,<br>SDHD,<br>ZBTB16,<br>PAFAH1B2,<br>NUMA1,<br>PICALM, EED | 3 | 0.11 |
| 15q | Gain | B2M,<br>HMGN2P46,<br>USP8,<br>MYO5A,<br>C15orf65,<br>TCF12,<br>MAP2K1,<br>SMAD3, PML,<br>NTRK3,<br>POLG, IDH2,<br>CRTC3, BLM,<br>FES, CHD2,<br>NUTM1,<br>BUB1B,<br>KNSTRN,<br>KNL1 | 3 | 0.11 |
| 17p | Loss | TP53, PER1,<br>GAS7,<br>MAP2K4,<br>NCOR1,<br>YWHAE,<br>USP6,<br>RABEP1,<br>FLCN,<br>SPECC1 | 3 | 0.11 |
| 6q | Loss | EPHA7,<br>CCDC88A,<br>PRDM1,<br>FOXO3,<br>ROS1, GOPC,<br>RSPO3,<br>PTPRK, SGK1,<br>MYB, BCLAF1,<br>TNFAIP3,<br>ECT2, LATS1,<br>ESR1,<br>ARID1B, EZR,<br>QKI,<br>FGFR1OP,<br>AFDN | 3 | 0.11 |
| 8p | Loss | PCM1,<br>ARHGEF10,<br>LEPROTL1,<br>WRN, NRG1,<br>NSD3, FGFR1,<br>ANK1, KAT6A,<br>IKBKB,<br>HOOK3 | 3 | 0.11 |
| 10p | Loss | LARP4B,<br>KLF6, GATA3,<br>MLLT10 | 2 | 0.07 |
| 14q | Loss | CCNB1IP1,<br>ARHGAP5,<br>BAZ1A,<br>NKX2-1,<br>FOXA1,<br>TRIP11,<br>GOLGA5 | 2 | 0.07 |
| 16q | Loss | CYLD,<br>HERPUD1,<br>CDH11, CBFB,<br>CTCF, CDH1,<br>ZFHX3,<br>RFWD3, MAF,<br>CBFA2T3,<br>FANCA | 2 | 0.07 |
| 8q | Gain | TCEAL1,<br>PLAG1,<br>CHCHD7,<br>PREX2,<br>NCOA2,<br>HEY1, CNBD1,<br>NBN,<br>RUNX1T1,<br>CDH17,<br>COX6C,<br>PABPC1,<br>UBR5,<br>RSPO2,<br>EIF3E,<br>CSMD3,<br>RAD21, EXT1,<br>MYC, NDRG1,<br>FAM135B,<br>RECQL4 | 2 | 0.07 |

### Aggregated Somatic Variants and Gene Specific Results

The results across analyses of SNVs and indels, structural variants (SV), and copy number variants (CNV) in MCL patients were aggregated to comprehensively describe the spectrum of mutations (Figure 2). The most commonly mutated gene among our cohort was *CCND1*, which consisted entirely of missense variants and translocations. The majority of the translocations in *CCND1* were the canonical t(11;14) translocation with IGH, however, two samples also harbored an intrachromosomal inversion that impacted *CCND1* (Supplemental Figure 7). All missense variants in *CCND1* were found in exon 1 of the gene in the N-terminal domain, and there was a mutational hotspot at amino acid position 36, where we identified 5 patients with mutations at this position, which is reported previously ^37^(Supplemental Figure 4). Conversely, all translocations in *CCND1* involved breakpoints located in the 5’ untranslated region of the gene, which is consistent with the nature of the t(11;14) translocation.

**Figure 2:**
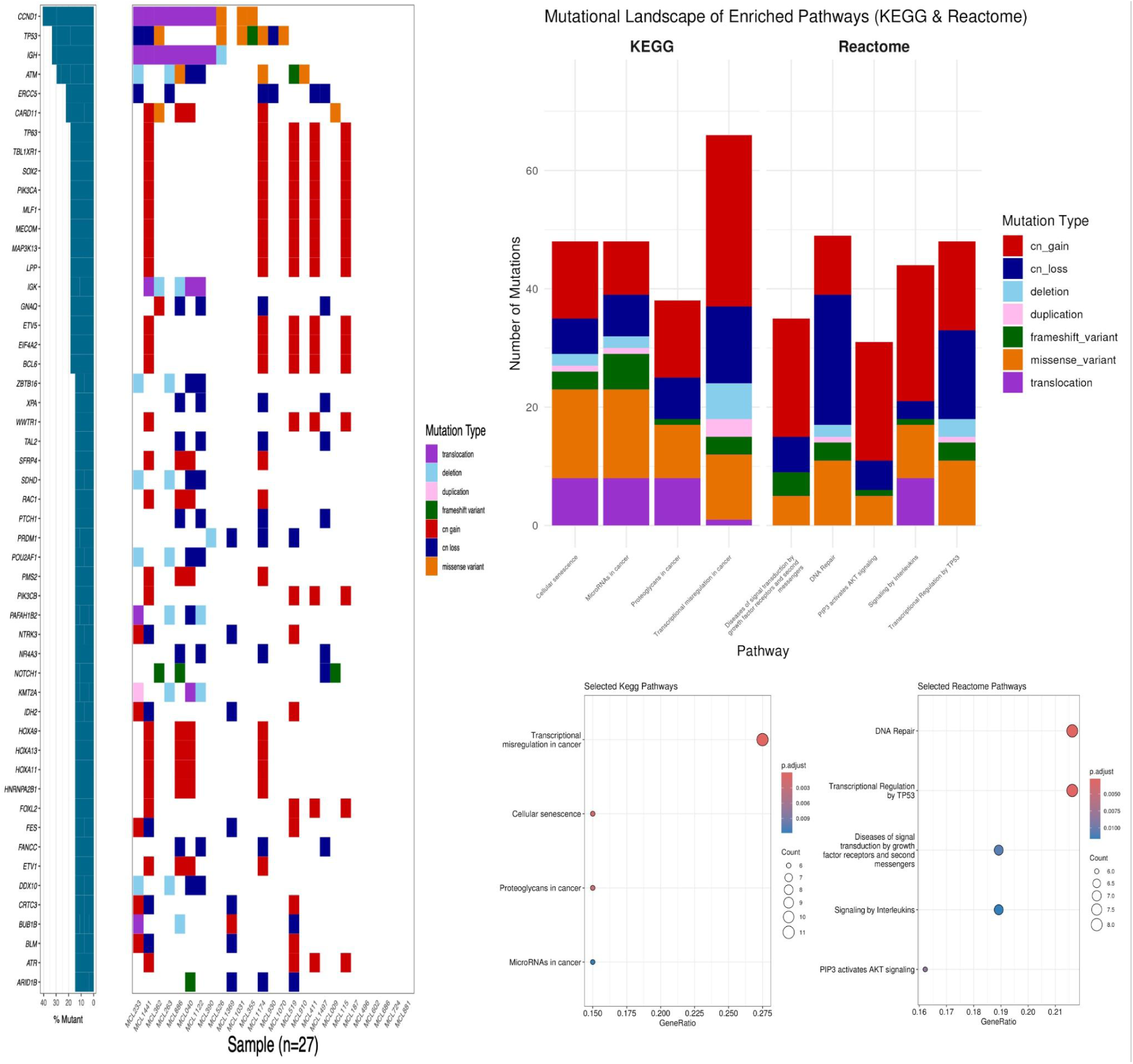
Aggregated Mutation Summary and Pathway-Level Alteration Analysis (a) Aggregated mutation landscape (waterfall plot) displaying the distribution of somatic mutations across samples. The frequency and type of mutations affecting genes with SNV, INDEL, copy number, or structural variant mutations in 4 or more patients. Mutated genes are shown in each row. Each column represents a patient in the cohort (n=27). The bar graph on the right summarizes the frequency of mutations for that gene across the entire cohort. For genes with multiple mutations in a single patient, only 1 mutation type is shown prioritizing the most severe mutation (listed in order in the legend). (b) Stacked bar plot summarizing the total number and types of mutations identified per pathway. Each bar represents a distinct biological pathway, and segments within each bar correspond to different mutation types, including copy number gains, copy number losses, point mutations (e.g., missense and frameshift), translocations, and duplications.(c) Pathway enrichment analysis (KEGG and Reactome). Each dot represents an enriched pathway. The dot size reflects the number of mutated genes within that pathway, while the color intensity corresponds to the statistical significance of enrichment (adjusted p-value), with darker shades indicating higher significance.

We observed the greatest diversity of variant types at *ATM*. At this locus we observed missense variants, frameshift variants, structural variants, and copy number losses. The SNVs and indels in *ATM* were spread out across the gene without a single hotspot. In this study *ATM* was often involved in large-scale deletions detected on the 11q chromosome arm. *TP53* also was mutated in multiple ways. The mutations we observed consisted of missense variants, frameshift variants, and copy number losses. Missense variants exclusively occurred in the p53 DNA-binding domain (Supplemental Figure 4). Interestingly, there was no overlap between the samples harboring SNVs and indels and the samples harboring copy number losses of TP53, suggesting that one mutational hit at *TP53* may be sufficient to impact MCL pathogenesis. SNVs and indels in *NOTCH1* are frequently found in MCL studies^38^. The frequency of *NOTCH1* variants in this study is similar to previous studies, and interestingly all 3 variants were frameshift mutations in the PEST domain and are potentially loss-of-function. The significance of these specific *NOTCH1* variants has been characterized in both MCL and other cancers, and have been associated with poor prognosis. The four patients harboring *NOTCH1* variants or copy loss (three at pre-treatment [PT] and one at second relapse) in this cohort had overall survival times of 149.65, 103.86, 64.87, and 32.45 months, respectively. Although *NOTCH1* alterations have previously been associated with adverse clinical outcomes, all four patients were alive at last follow-up which is not uniformly suggestive of poor prognosis, highlighting the need for larger cohorts.

### Aggregated Pathway Analysis

#### Aggregated Pathway Analysis

Pathway enrichment analysis identified significant enrichment of biological processes related to transcriptional regulation, DNA damage response, oncogenic signaling, cellular senescence, and immune signaling (Figure 2). Reactome pathways included DNA repair, signaling by interleukins, transcriptional regulation by *TP53*, *PIP3* activates *AKT* signaling, and diseases of signal transduction by growth factor receptors and second messengers. Significant KEGG pathways included transcriptional misregulation in cancer, cellular senescence, microRNAs in cancer, and proteoglycans in cancer.

These pathways were affected by multiple classes of somatic alterations, including SNVs, indels, structural variants, copy-number gains, and copy-number losses.

Copy-number alterations contributed substantially to several enriched pathways. Recurrent copy-number gains were most prominent in transcriptional misregulation in cancer, *PIP3* activates *AKT* signaling, and signaling by interleukins, affecting genes including *BCL6, ETV1, HOXA9, HOXA11, PIK3CA, PIK3CB, SOX2, and HNRNPA2B1*.

In contrast, copy-number losses were most prominent in the DNA repair pathway, where recurrent deletions affected genes including *ATM* and *ERCC5* (Figure 2b). These observations indicate that multiple classes of somatic alterations converge on a limited number of biologically relevant pathways in MCL. Because copy-number alterations were summarized at the chromosomal arm level, the contribution of individual genes within these regions should be interpreted cautiously.

#### Transcriptional Misregulation in Cancer

This pathway, as annotated in the KEGG database, includes 201 genes. Of the 50 genes mutated recurrently in our cohort, 11 were part of this pathway, representing 5.5% of the total pathway genes (11/201). The transcriptional misregulation pathway exhibited the highest number of mutated genes among all pathways we interrogated (Supplemental Table 6). We observed a total of 66 mutations across these 11 genes. Importantly, copy number gains were the most frequent event. Key transcriptional regulators such as *ETV1*, *HOXA9*, *HOXA11*, *MLF1*, *ETV5*, *BCL6*, *ZBTB16*, and *KMT2A* were recurrently amplified. These genes regulate diverse biological processes, including B-cell differentiation (*BCL6*), hematopoietic development (*HOXA9*, *HOXA11*, *MLF1*), transcriptional regulation (*ETV1*, *ETV5*, *ZBTB16*), and chromatin remodeling (*KMT2A*), suggesting that multiple transcriptional programs may be disrupted in MCL rather than a single common regulatory mechanism.

#### DNA Repair

In the DNA repair pathway, we identified 8 mutated genes which constituted 2.3% of the total pathway genes (8/335), accounting for 49 distinct mutations. The most notable finding was the alteration of *ERCC5,* which is a gene responsible for a nucleotide excision repair protein, which was found to be mutated in 6 patients, all due to copy number losses, suggesting compromised nucleotide excision repair.

#### PIP3/AKT Signaling

The PIP3/AKT pathway, which plays a critical role in cellular survival, proliferation, and resistance to apoptosis^39^, harbored 31 mutations affecting 6 genes, representing 2.2% of the 273 genes in the pathway, including *PIK3CA*, *PIK3CB*, and other PI3K-pathway members. Notably, 5 out of 6 genes (83%) showed copy number gains, indicating pathway activation through genomic amplification. These alterations may promote tumorigenesis through activation of pro-survival and proliferative signaling pathways, particularly the PI3K/AKT axis, which regulates cell growth, metabolism, and apoptosis. In MCL, PI3K signaling has a central pathogenic role, with evidence that lymph node–resident MCL cells exhibit aberrant expression of the PI3Kγ isoform, contributing to enhanced proliferation, survival, and migration^40^. Dual inhibition of PI3Kδ/γ has been shown to more effectively suppress MCL cell growth and tumor progression compared to isoform-selective approaches, underscoring the functional importance of this pathway in disease biology and therapeutic resistance.^41^

#### Signaling by Interleukins

This immune signaling pathway exhibited mutations in 7 genes, representing 1.5% of the 473 pathway genes (7/473), with a total of 44 mutation events, 23 of which were copy number gains. Notably, key genes such as *SOX2*, *PIK3CA*, *BCL6*, *PIK3CB*, and *HNRNPA2B1* showed copy number gains.

#### Transcriptional Regulation by TP53

The Reactome transcriptional regulation by TP53 pathway contained 9 altered genes, representing 2.1% of the 433 genes annotated within the pathway. A total of 48 alteration events were identified, involving genes such as *TP53, ATM, PRDM1, FANCC, BLM, PMS2, ATR, and BCL6.* Alterations included copy number gains, copy number losses, missense mutations, frameshift mutations, and structural variants.

### Recurrent RNA Fusions Events

Analysis of RNA fusions in 8 RNA-Seq samples revealed recurrent non-read through fusions (Figure 3), with the most common gene pairs being KCNQ1OT1-TRIM68 (3/8; 37.5%), EML2-NOVA2, (3/8; 37.5%), SWT1-RNF2 (3/8; 37.5%), CDC42EP4-HNRNPU (2/8; 25.0%), REV3L-CENPW (2/8; 25.0%), SEPTIN7P2-PSPH (2/8; 25.0%), URI1-VSTM2B (2/8; 25.0%), VTI1B-PLEK2 (2/8; 25.0%). All other non read-through detected RNA fusions are summarized in Supplemental Table 7. We asked if any of the gene fusion pairs are present in the Cancer Gene Census (CGC). Among the identified fusion events, the gene *CIC*, which was observed in a fusion pair *MEGF8* in one sample, was the only gene present in the CGC. The CIC::MEGF8 fusion, previously reported in melanoma and has been suggested to cause transcriptional dysregulation through CIC loss of function^42^. Furthermore, an analysis of the genomic regions within 10 kb upstream or downstream of the fusion breakpoints identified *MED12*, a CGC-listed gene, located near the *NLGN3::COL25A1* fusion, although *NLGN3* itself is not part of the CGC.

**Figure 3:**
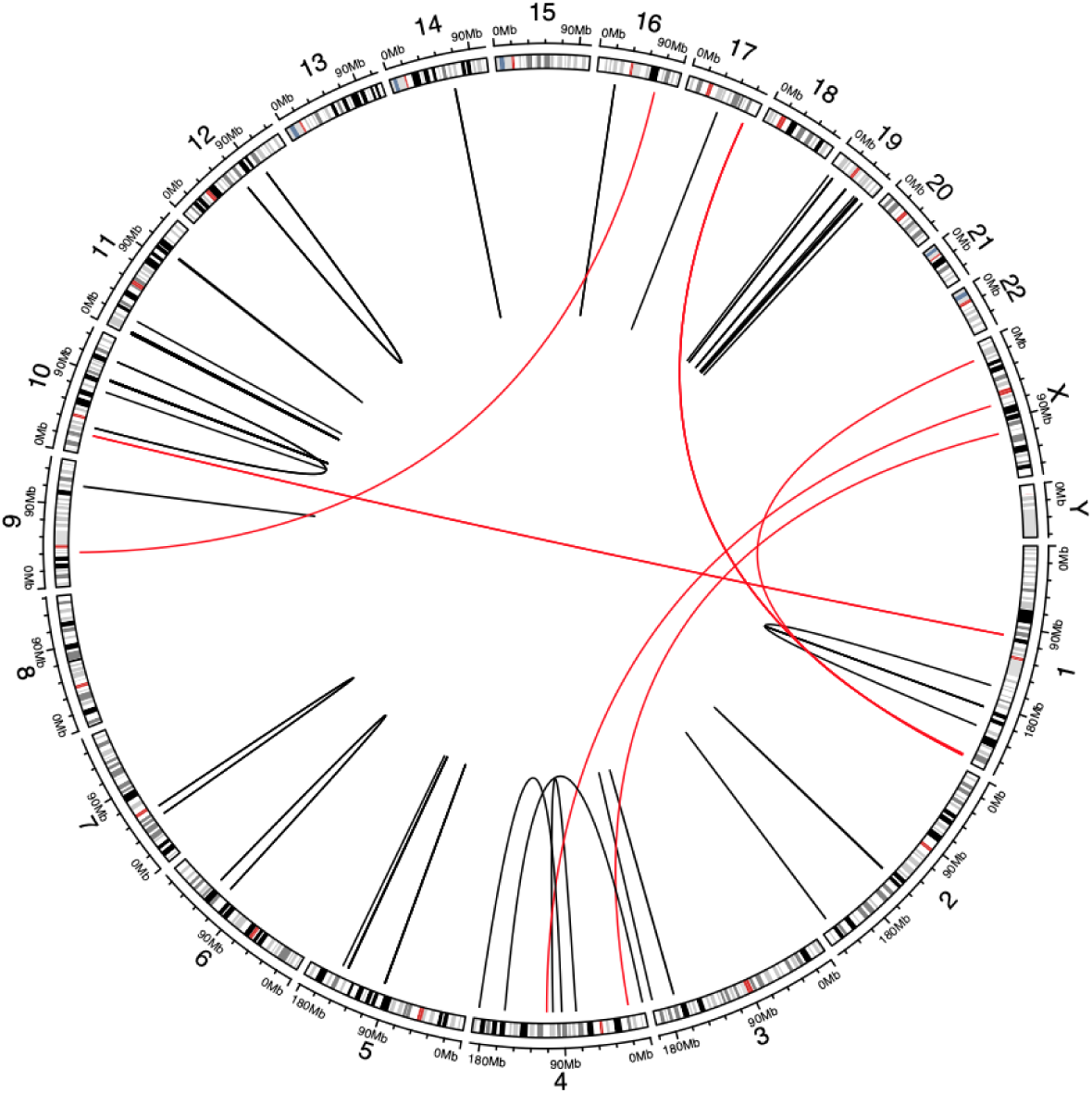
Non-readthrough RNA fusion transcripts detected in the 8 RNASeq MCL samples. Interchromosomal fusions (2 different chromosomes) are depicted with the red lines, while intrachromosomal fusions (within the same chromosome) are depicted with the black lines.

## Discussion

This study provides an integrated genomic and transcriptomic characterization of mantle cell lymphoma (MCL), highlighting recurrent genomic alterations and the biological pathways that may contribute to disease heterogeneity and progression. MCL is an aggressive and clinically heterogeneous B-cell malignancy driven by a variety of genetic aberrations. The canonical t(11;14)(q13;q32) translocation that facilitates the overexpression of *CCND1* defines the disease, but additional mutations and chromosomal alterations influence its progression and clinical outcomes. In this study, we conducted a comprehensive genomic and transcriptomic analysis integrating WES, WGS, and RNA-seq across 27 patient samples. This multi-platform design enabled simultaneous characterization of SNVs, indels, SVs, CNAs, and RNA fusion events, providing a detailed view of the somatic landscape of MCL. Our analysis confirmed the presence of established driver genes including *ATM*, *TP53*, and *CCND1*,^5^ ^43^ ^44^ and revealed novel candidate genes including *ASXL1*. Because *ASXL1* is a gene frequently associated with clonal hematopoiesis of indeterminate potential (CHIP), we interpreted these findings with caution. The variants were identified primarily in nodal tissue, where contamination by circulating CHIP clones is expected to be lower than in peripheral blood or bone marrow. Nevertheless, the contribution of a coexisting hematopoietic clone cannot be excluded.

Disruption of the DNA damage response emerged as one of the predominant genomic features of our cohort, with recurrent alterations affecting the key DDR genes ATM and TP53. Alterations in *ATM* and *TP53* were among the most frequent events in the cohort (Figure 2), supporting recurrent involvement of DNA damage response (DDR) genes in MCL. *ATM* was altered by multiple variant classes, including missense variants, frameshift variants, and large-scale copy-number losses on chromosome 11q, whereas *TP53* was affected by SNV/indel events and copy-number losses on chromosome 17p. When SNV/indel and copy-number events were considered together, *ATM* and *TP53* alterations showed limited overlap, with most affected samples harboring an alteration of only one of these two genes. This pattern suggests that disruption of DDR-related biology may occur through different genomic mechanisms across tumors. Furthermore, there were 3 patients with relatively short survival times (19 months, 1 month, 44 months) who had mutations in either *TP53* or *ATM*, the shortest being a copy loss in *TP53*. However, because two patients harbored alterations affecting both *ATM* and *TP53*, and because no mutually exclusive gene pairs reached statistical significance, these observations should be interpreted descriptively rather than as evidence of formal mutual exclusivity or clear clinical associations.

Structural variant analysis revealed recurrent chromosomal rearrangements that extend beyond the canonical CCND1 translocation and further define the genomic architecture of MCL. Structural variant analysis provided insight into large-scale genomic rearrangements that shape the MCL genome. Whole-genome sequencing revealed 142 structural events across 10 samples. The hallmark t(11;14)(q13;q32) translocation was detected in eight samples. In addition to this canonical event, we identified recurrent rearrangements involving *KMT2A* and *PAFAH1B2.* Rearrangements of *KMT2A*, identified as structural alterations in 4 of 8 patients, suggesting that alterations affecting this gene may represent recurrent secondary genomic events in MCL. As KMT2A is a key regulator of chromatin state and transcriptional programs, disruption of this locus could have functional consequences; however, additional studies will be required to determine its role in MCL biology and clinical outcome. Among the four patients with *KMT2A* rearrangements, two received four lines of treatment and had overall survival times of 44 and 99 months. The remaining two patients did not progress after frontline treatment and had overall survival times of 139 and 92 months. Given these mixed clinical outcomes and the small number of cases, the present data do not support a clear association between *KMT2A* rearrangement and poor clinical outcome; larger cohorts will be required to determine its clinical significance. These results highlight the importance of incorporating WGS for comprehensive detection of structural variants and for understanding the full spectrum of genomic alterations in MCL. This result is further supported by a recent case of classic MCL in which a complex t(6;11;14) rearrangement involving *KMT2A* was identified alongside CCND1::IGH, with *KMT2A* rearrangement present in a substantial proportion of tumor cells, underscoring its potential role as a recurrent, clonally relevant secondary event in MCL pathogenesis. ^45^

Copy-number alterations represented another major source of genomic disruption in MCL, affecting multiple chromosomal regions involved in tumor suppressor function and oncogenic signaling. Copy-number analysis identified recurrent chromosomal arm-level events, with losses occurring more frequently than gains consistent with previous work^36^. Recurrent deletions were observed on 17p, 11q, and 13q which could impact *TP53*, *ATM,* and *ERCC5* respectively, further implicating defects in DDR. Gains were most frequent on 3q, 7p, and 15q, affecting genes such as *BCL6*, *HOXA9*, and *ETV1*, which regulate transcription and differentiation. These patterns align with previous reports in MCL but provide enhanced resolution when considered alongside SNV and SV data. Additionally, amplification at transcriptional regulators such as *BCL6* and *HOXA9* suggests increased gene dosage, which has been previously reported in B cell lymphomas^46^ and has been shown/suggested to enhance oncogenic signaling.

Beyond established MCL driver genes, our integrated sequencing approach identified several candidate genes that may contribute to disease biology and warrant further investigation. The integration of multiple sequencing platforms enabled identification of several novel recurrently mutated genes, (*ASXL1*, *TNC*, *PCDH15*, and *ZNF804B)*.

Among these, *ASXL1* was the only gene to reach nominal significance in the dN/dScv analysis, suggesting evidence of positive selection. *ASXL1* encodes a chromatin-associated protein involved in histone modification, and its mutation has been linked to poor prognosis in myeloid malignancies ^47^ . The presence of *ASLX1* mutations in MCL suggests a similar mechanism of altered chromatin regulation that could influence transcriptional programs in MCL.

Although individual genomic alterations varied substantially between patients, pathway-level analysis demonstrated convergence on a limited number of recurrent biological processes. To contextualize the biological significance of the diverse somatic alterations identified across variant classes, we performed an integrated pathway enrichment analysis using the combined set of genes affected by single-nucleotide variants, indels, structural variants, and copy number alterations. Despite marked inter-patient heterogeneity, mutations across the cohort were non-randomly concentrated in a limited set of functional networks (Figure 2b–c). The most significantly enriched pathways (q <0.05) notably included transcriptional misregulation in cancer, DNA repair, PI3K/AKT signaling, and signaling by interleukins. The significantly enriched pathways were disrupted by multiple classes of genomic lesions (e.g. copy-number gains and losses, structural translocations, and point mutations).

Within the transcriptional misregulation pathway (KEGG), recurrent amplifications were observed in transcriptional regulators such as *BCL6*, *ETV1*, *HOXA9*, *HOXA11*, and *KMT2A*, suggesting aberrant control of cell identity and differentiation programs. DNA repair genes including *ATM*, *ERCC5* were frequently inactivated through point mutations and 11q deletions. The PI3K/AKT signaling network was predominantly altered through copy-number gains of *PIK3CA*, *PIK3CB*, and *AKT1*, implicating enhanced prosurvival signaling. Constitutive activation of the PI3K/AKT pathway, potentially driven by *PTEN* loss, has been previously identified in aggressive blastoid mantle cell lymphoma and is proposed to contribute to its pathogenesis.^48^ Finally, enrichment of interleukin signaling and NF-κB–associated pathways was driven by CNA gains in *REL*, *BCL2*, and *NFKBIE*, consistent with immune–oncogenic cross-talk and chronic signaling activation in MCL.^49^

Together, these findings indicate that diverse classes of somatic alterations in MCL converge on a limited set of biological pathways, including transcriptional regulation, DNA damage response, PI3K/AKT signaling, and immune-related signaling networks. Integrating mutations, structural variants, and copy-number alterations at the pathway level helped contextualize the functional relevance of recurrent genomic events identified across patients. These recurrently affected pathways may represent biologically important processes in MCL and warrant further investigation as potential therapeutic targets.

Gene-level significance analysis identified TP53 as the only statistically significant driver, while additional recurrently mutated genes provided complementary biological insights despite not meeting significance thresholds.Although *TP53* was the only gene reaching statistical significance in dN/dScv, the broader mutation set included alterations in genes linked to B-cell receptor and NF-κB signaling, including *CD79B*, *CARD11*, and *PLCG2*. These genes did not meet recurrence or FDR-based significance thresholds in this cohort, so they should be interpreted cautiously; however, their presence supports the pathway-level observation that immune and B-cell signaling programs are recurrently affected across multiple classes of somatic alteration.

## Limitations and Future Directions

The main limitation of this study is the relatively small cohort size, which constrains statistical power for detecting rare mutations and interactions. The low overall mutation burden in MCL further limits discovery of mutually exclusive events. In addition, RNA-seq was available for only 8 samples, which restricts detection of fusion transcripts. Despite these constraints, the integration of WES, WGS, and RNA-seq provides a strong foundation for defining MCL’s somatic mutation architecture.

Future studies should aim to validate novel candidate mutations using functional assays. CRISPR-engineered models and drug-sensitivity profiling will be valuable tools to determine whether alterations in *ASXL1*, and other genes known to impact pathogenesis in others cancers, but were observed at a low-prevalence in this study.

Expanding the cohort size will also allow more robust association of genomic profiles with clinical outcomes, facilitating the development of mutation-based prognostic markers.

## Conclusions

This work demonstrates that integration of WES, WGS, and RNA-seq provides a strong framework for characterizing the somatic alteration landscape of MCL. Our findings support the recurrent involvement of DDR genes, particularly *ATM* and *TP53*, and highlight copy-number alterations as an additional mechanism contributing to disruption of key tumor suppressor and signaling pathways. The detection of recurrent structural rearrangements, including events involving *CCND1*::*IGH* and *KMT2A*, further illustrates the value of WGS for capturing alterations not fully resolved by exome sequencing alone. Finally, pathway-level integration helped consolidate diverse genomic findings into recurrently affected biological processes, including DNA repair, transcriptional regulation, PI3K/AKT signaling, and immune-related signaling. Together, these results provide a comprehensive genomic description of this MCL cohort and identify candidate alterations and pathways that warrant further functional and clinical investigation.

## Data Availability

The data supporting the findings of this study are provided within the manuscript and its Supplementary Information. Additional data are available from the corresponding authors upon reasonable request.

## Authorship Contributions

B.K. and O.L.G conceptualized the study. M.G., O.L.G., B.K. and T.A.F. developed the experimental design with assistance from B.L., F.G.. B.K., D.R-G, and T.A.F. acquired samples and clinical data. B.L., C.N.J., M.M., Z.S., K.C., F.G., M.G., and O.L.G. performed data analysis. B.L., C.N.J., M.M., A.D., F.G., T.A.F., M.G., and O.L.G. prepared figures and tables. B.L., C.N.J., F.G., O.L.G., M.G. and T.A.F. wrote the manuscript with assistance from D.R-G. All authors approved the final version of the manuscript.

## Acknowledgements

The authors thank Alvin J. Siteman Cancer Center at Washington University School of Medicine and Barnes-Jewish Hospital and the Institute of Clinical and Translational Sciences (ICTS) at Washington University, for the use of the Tissue Procurement Core, which provided DNA and RNA isolation services.

The Siteman Cancer Center is supported in part by an NCI Cancer Center Support Grant #P30 CA091842 and the ICTS is funded by the National Institutes of Health’s NCATS Clinical and Translational Science Award (CTSA) program grant #UL1 TR002345.

M.G. was supported by a National Institutes of Health, National Human Genome Research Institute grant (K99 HG007940), a National Cancer Institute grant (U01 CA248235) and a V Scholar Award from the V Foundation for Cancer Research (Award: V2018-007)

This work was supported by the Siteman Investment Program Team Science Award (T.A.F, B.K.), the Larry and Winnie Chiang Lymphoma Fellowship (T.A.F., F.G.) and Blood Cancer United Specialized Center of Research (SCOR). (O.L.G., M.G., T.A.F.,B.K.)

T.A.F is supported by NIH P50CA171063, P30CA91842, Blood Cancer United Specialized Center of Research (SCOR), the Paula and Rodger Riney Foundation, and the Steinberg Family Cancer Fund and the Siteman Cancer Center Research Development Award.

FG is supported by The Paul Calabresi Career Development Award (K12 5K12CA167540-07; F.G.). FG and CNJ were supported by the National Institutes of Health, National Cancer Institute grant (K22 CA266743) and the Marge and Charles J Schott Foundation Blood Cancer Research fund

## References

[1] Jares P, Colomer D, Campo E. Molecular pathogenesis of mantle cell lymphoma. J Clin Invest. 2012;122(10):3416–3423.

[2] Yin CC, Luthra R. Molecular detection of t(11;14)(q13;q32) in mantle cell lymphoma. Methods Mol Biol. 2013;999:211–216.

3. [3] Mantle cell lymphoma with a novel t(11;12)(q13;p11.2): a proposed alternative mechanism of CCND1 up-regulation. Human Pathology. 2017;64:207-212.

[4] Yin CC, Luthra R. Molecular detection of t(14;18)(q32;q21) in follicular lymphoma. Methods Mol Biol. 2013;999:203–209.

[5] Beà S, Valdés-Mas R, Navarro A, Salaverria I, Martín-Garcia D, Jares P, et al. Landscape of somatic mutations and clonal evolution in mantle cell lymphoma. Proc Natl Acad Sci U S A. 2013;110(45):18250–18255.

[6] Kridel R, Chan FC, Mottok A, Boyle M, Farinha P, Tan K, et al. Histological Transformation and Progression in Follicular Lymphoma: A Clonal Evolution Study. PLoS Med. 2016;13(12):e1002197.

[7] Hill HA, Qi X, Jain P, Nomie K, Wang Y, Zhou S, et al. Genetic mutations and features of mantle cell lymphoma: a systematic review and meta-analysis. Blood Adv. 2020;4(13):2927–2938.

[8] Dreyling M, Doorduijn J, Giné E, Jerkeman M, Walewski J, Hutchings M, et al. Addition of autologous stem-cell transplantation to an ibrutinib-containing first-line treatment in patients aged 18-65 years with mantle cell lymphoma (TRIANGLE): 4·5-year follow-up of a three-arm, randomised, open-label, phase 3 superiority trial of the European MCL Network. Lancet. 2026;407(10542):1953–1967.

[9] Parekh S, Weniger MA, Wiestner A. New molecular targets in mantle cell lymphoma. Semin Cancer Biol. 2011;21(5):335–346.

[10] Kumar A, Soumerai J, Abramson JS, Barnes JA, Caron P, Chhabra S, et al. Zanubrutinib, obinutuzumab, and venetoclax for first-line treatment of mantle cell lymphoma with a TP53 mutation. Blood. 2025;145(5):497–507.

[11] Griffith M, Griffith OL, Smith SM, Ramu A, Callaway MB, Brummett AM, et al. Genome Modeling System: A Knowledge Management Platform for Genomics. PLOS Computational Biology. 2015;11(7):e1004274.

12. [12] Li H. Aligning sequence reads, clone sequences and assembly contigs with BWA-MEM. *arXiv preprint arXiv:13033997*. Published online 2013. http://arxiv.org/abs/1303.3997

[13] Faust GG, Hall IM. SAMBLASTER: fast duplicate marking and structural variant read extraction. Bioinformatics. 2014;30(17):2503–2505.

14. [14] Artem Tarasov PP. sambamba. Published online 2012. http://lomereiter.github.io/sambamba/docs/sambamba-view.html

[15] Li H, Handsaker B, Wysoker A, Fennell T, Ruan J, Homer N, et al. The Sequence Alignment/Map format and SAMtools. Bioinformatics. 2009;25(16):2078–2079.

[16] Larson DE, Harris CC, Chen K, Koboldt DC, Abbott TE, Dooling DJ, et al. SomaticSniper: identification of somatic point mutations in whole genome sequencing data. Bioinformatics. 2012;28(3):311–317.

[17] Koboldt DC, Zhang Q, Larson DE, Shen D, McLellan MD, Lin L, et al. VarScan 2: somatic mutation and copy number alteration discovery in cancer by exome sequencing. Genome Res. 2012;22(3):568–576.

[18] Cibulskis K, Lawrence MS, Carter SL, Sivachenko A, Jaffe D, Sougnez C, et al. Sensitive detection of somatic point mutations in impure and heterogeneous cancer samples. Nat Biotechnol. 2013;31(3):213–219.

[19] Saunders CT, Wong WSW, Swamy S, Becq J, Murray LJ, Cheetham RK. Strelka: accurate somatic small-variant calling from sequenced tumor-normal sample pairs. Bioinformatics. 2012;28(14):1811–1817.

[20] Ye K, Schulz MH, Long Q, Apweiler R, Ning Z. Pindel: a pattern growth approach to detect break points of large deletions and medium sized insertions from paired-end short reads. Bioinformatics. 2009;25(21):2865–2871.

[21] McKenna A, Hanna M, Banks E, Sivachenko A, Cibulskis K, Kernytsky A, et al. The Genome Analysis Toolkit: a MapReduce framework for analyzing next-generation DNA sequencing data. Genome Res. 2010;20(9):1297–1303.

[22] Barnell EK, Ronning P, Campbell KM, Krysiak K, Ainscough BJ, Sheta LM, et al. Standard operating procedure for somatic variant refinement of sequencing data with paired tumor and normal samples. Genetics in medicine : official journal of the American College of Medical Genetics. 2019;21(4). doi:10.1038/s41436-018-0278-z

[23] Chen X, Schulz-Trieglaff O, Shaw R, Barnes B, Schlesinger F, Källberg M, et al. Manta: rapid detection of structural variants and indels for germline and cancer sequencing applications. Bioinformatics. 2016;32(8):1220-1222.

[24] Spies N, Zook JM, Salit M, Sidow A. svviz: a read viewer for validating structural variants. Bioinformatics. 2015;31(24):3994–3996.

[25] Haas BJ, Dobin A, Ghandi M, Van Arsdale A, Tickle T, Robinson JT, et al. Targeted characterization of fusion transcripts in tumor and normal tissues via FusionInspector. Cell Rep Methods. 2023;3(5):100467.

26. [26] clusterProfiler 4.0: A universal enrichment tool for interpreting omics data. The Innovation. 2021;2(3):100141.

[27] Luo W, Friedman MS, Shedden K, Hankenson KD, Woolf PJ. GAGE: generally applicable gene set enrichment for pathway analysis. BMC Bioinformatics. 2009;10:161.

[28] Luo W, Brouwer C. Pathview: an R/Bioconductor package for pathway-based data integration and visualization. Bioinformatics. 2013;29(14):1830–1831.

[29] Yu G, He QY. ReactomePA: an R/Bioconductor package for reactome pathway analysis and visualization. Mol Biosyst. 2016;12(2):477–479.

[30] Pedersen BS, Bhetariya PJ, Brown J, Kravitz SN, Marth G, Jensen RL, et al. Somalier: rapid relatedness estimation for cancer and germline studies using efficient genome sketches. Genome Medicine. 2020;12(1):1–9.

[31] Gelsi-Boyer V, Trouplin V, Roquain J, Adélaïde J, Carbuccia N, Esterni B, et al. ASXL1 mutation is associated with poor prognosis and acute transformation in chronic myelomonocytic leukaemia. British Journal of Haematology. 2010;151(4):365–375.

[32] Boultwood J, Perry J, Zaman R, Fernandez-Santamaria C, Littlewood T, Kusec R, et al. High-density single nucleotide polymorphism array analysis and ASXL1 gene mutation screening in chronic myeloid leukemia during disease progression. Leukemia. 2010;24(6):1139–1145.

[33] Jain P, Wang M. High-risk MCL: recognition and treatment. Blood. 2025;145(7):683–695.

[34] Pararajalingam P, Coyle KM, Arthur SE, Thomas N, Alcaide M, Meissner B, et al. Coding and noncoding drivers of mantle cell lymphoma identified through exome and genome sequencing. Blood. 2020;136(5). doi:10.1182/blood.2019002385

[35] Nadeu F, Martin-Garcia D, Clot G, Díaz-Navarro A, Duran-Ferrer M, Navarro A, et al. Genomic and epigenomic insights into the origin, pathogenesis, and clinical behavior of mantle cell lymphoma subtypes. Blood. 2020;136(12):1419–1432.

[36] Halldórsdóttir AM, Sander B, Göransson H, Isaksson A, Kimby E, Mansouri M, et al. High-resolution genomic screening in mantle cell lymphoma--specific changes correlate with genomic complexity, the proliferation signature and survival. Genes Chromosomes Cancer. 2011;50(2):113–121.

[37] Mohanty A, Sandoval N, Das M, Pillai R, Chen L, Chen RW, et al. CCND1 mutations increase protein stability and promote ibrutinib resistance in mantle cell lymphoma. Oncotarget. 2016;7(45). doi:10.18632/oncotarget.12434

[38] Kridel R, Meissner B, Rogic S, Boyle M, Telenius A, Woolcock B, et al. Whole transcriptome sequencing reveals recurrent NOTCH1 mutations in mantle cell lymphoma. Blood. 2012;119(9):1963–1971.

[39] Shin I, Yakes FM, Rojo F, Shin NY, Bakin AV, Baselga J, et al. PKB/Akt mediates cell-cycle progression by phosphorylation of p27(Kip1) at threonine 157 and modulation of its cellular localization. Nat Med. 2002;8(10):1145–1152.

[40] Jiang M, Zhang K, Zhang Z, Zeng X, Huang Z, Qin P, et al. PI3K/AKT/mTOR Axis in Cancer: From Pathogenesis to Treatment. MedComm (2020). 2025;6(8):e70295.

[41] Till KJ, Abdullah M, Alnassfan T, Janet GZ, Marks T, Coma S, et al. Roles of PI3Kγ and PI3Kδ in mantle cell lymphoma proliferation and migration contributing to efficacy of the PI3Kγ/δ inhibitor duvelisib. Sci Rep. 2023;13(1):3793.

[42] Hida T, Idogawa M, Sato S, Kiniwa Y, Kato J, Horimoto K, et al. Fusion Gene Detection in Driver Mutation-Negative Melanomas Using RNA-Based Anchored Multiplex Polymerase Chain Reaction. Pigment Cell & Melanoma Research. 2025;38(6):e70056.

[43] Greiner TC, Dasgupta C, Ho VV, Weisenburger DD, Smith LM, Lynch JC, et al. Mutation and genomic deletion status of ataxia telangiectasia mutated (ATM) and p53 confer specific gene expression profiles in mantle cell lymphoma. Proc Natl Acad Sci U S A. 2006;103(7):2352–2357.

[44] Zhang J, Jima D, Moffitt AB, Liu Q, Czader M, Hsi ED, et al. The genomic landscape of mantle cell lymphoma is related to the epigenetically determined chromatin state of normal B cells. Blood. 2014;123(19):2988–2996.

[45] Hu S, Tang G. Mantle cell lymphoma with KMT2A rearrangement. Blood. 2025;146(20):2491–2491.

[46] McLachlan T, Matthews WC, Jackson ER, Staudt DE, Douglas AM, Findlay IJ, et al. B-cell Lymphoma 6 (BCL6): From Master Regulator of Humoral Immunity to Oncogenic Driver in Pediatric Cancers. Mol Cancer Res. 2022;20(12):1711–1723.

[47] Bidikian A, Kantarjian H, Jabbour E, Short NJ, Patel K, Ravandi F, et al. Prognostic impact of ASXL1 mutations in chronic phase chronic myeloid leukemia. Blood Cancer J. 2022;12(10):144.

[48] Rudelius M, Pittaluga S, Nishizuka S, Pham THT, Fend F, Jaffe ES, et al. Constitutive activation of Akt contributes to the pathogenesis and survival of mantle cell lymphoma. Blood. 2006;108(5):1668–1676.

[49] Sun R, Wang J, Young KH. Oncogenic Signaling Pathways and Pathway-Based Therapeutic Biomarkers in Lymphoid Malignancies. Critical reviews in oncogenesis. 2017;22(5-6):527.

