## Supplemental Figures for "Surveying the Genomic Landscape of Mantle Cell Lymphoma Indicates the Importance of Multimodal Genomic and Transcriptomic Exploration"

List of supplemental data:

**Figures**

Supplemental figure 1: Sample related analysis using Somalier.

Supplemental Figure 2: Distribution of somatic variant consequences identified across the MCL cohort.

Supplemental figure 3: Landscape of recurrent somatic SNVs and indels in the MCL cohort.

Supplementary Figure 4. Protein-level distribution of recurrent somatic mutations in MCL.
Supplemental figure 5: Co-occurrence analysis of recurrent somatic SNVs and indels.

Supplementary Figure 6. Landscape of recurrent structural variants in the MCL cohort.

Supplementary Figure 7. Representative split-read evidence for somatic intrachromosomal inversions involving CCND1.

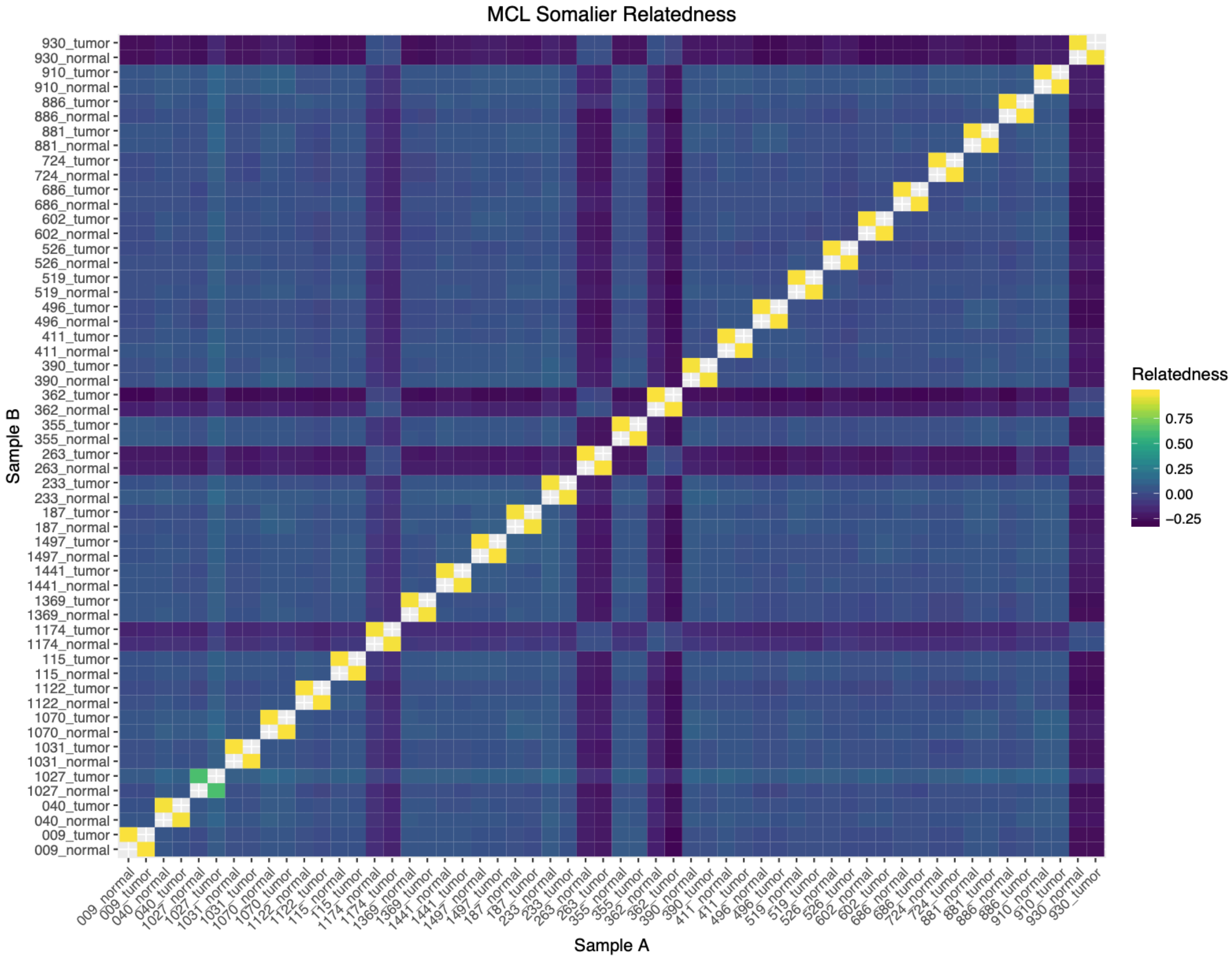


**Supplemental figure 1: Sample related analysis using Somalier.**

Heatmap of pairwise relatedness score generated with Somalier (Pedersen et al., 2020) for tumor and matched normal samples. Lower values indicate distant relatedness.

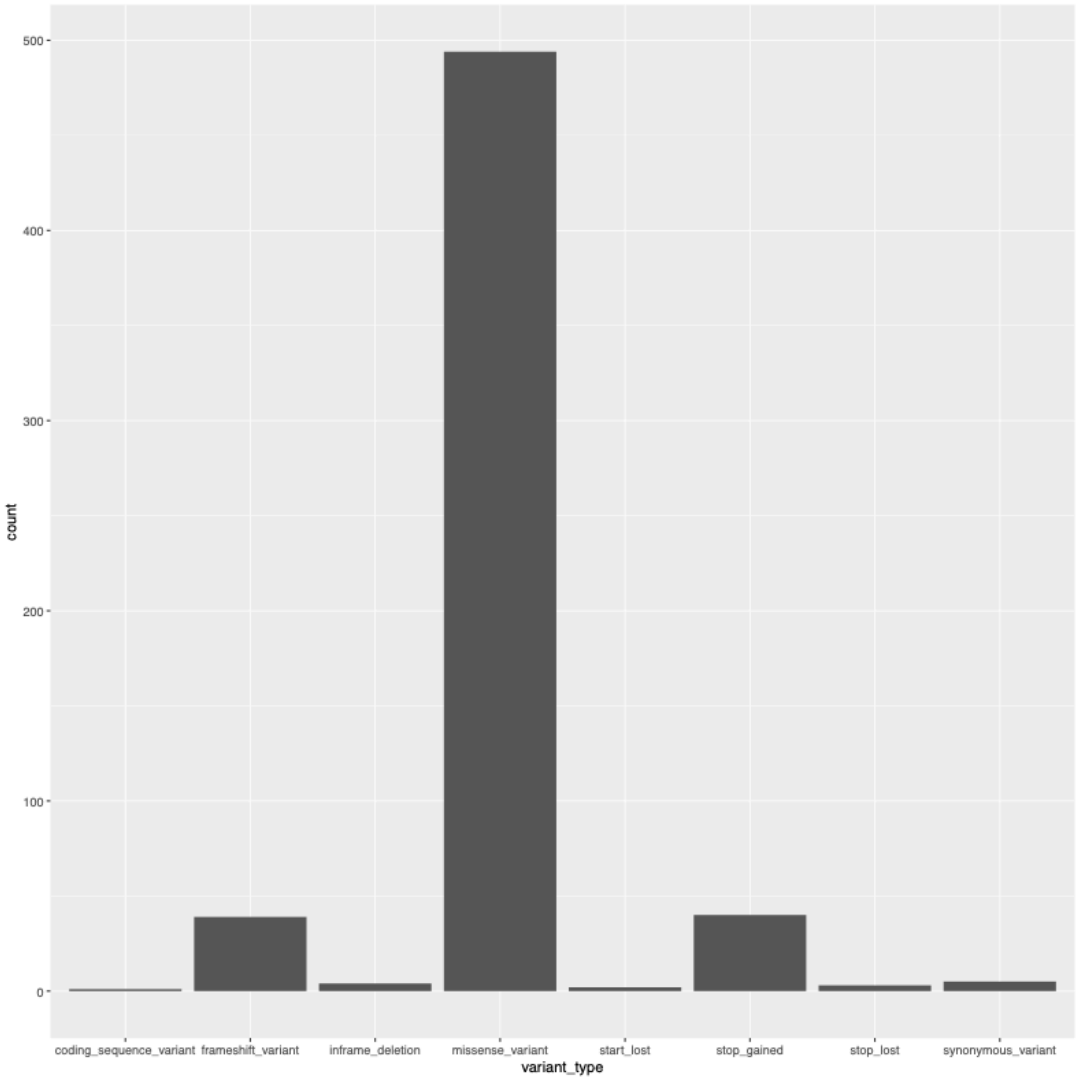


**Supplemental Figure 2: Distribution of somatic variant consequences identified across the MCL cohort.**

Bar plot showing the number of somatic variants classified by predicted functional consequence following variant annotation. Missense variants represent the predominant mutation class, followed by stop-gained and frameshift variants, whereas synonymous and splice-site variants were less frequent

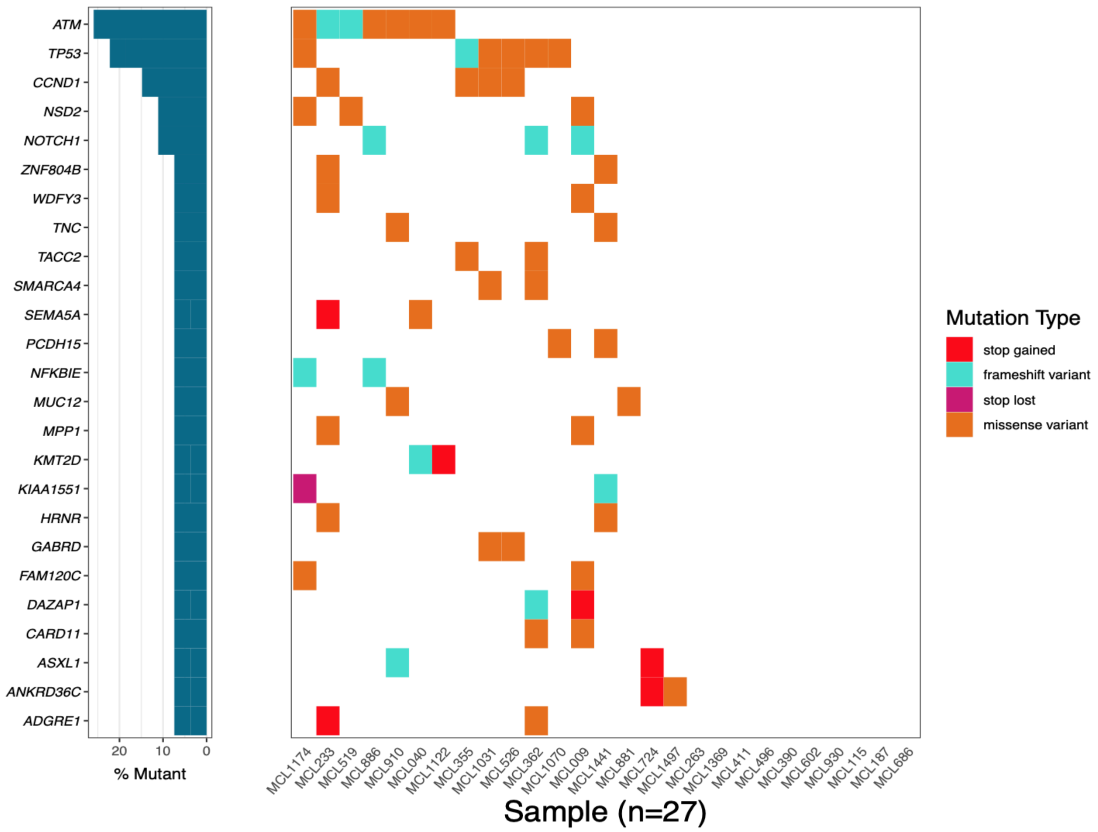


**Supplemental figure 3: Landscape of recurrent somatic SNVs and indels in the MCL cohort.**

Waterfall plot summarizing recurrent coding SNVs and indels identified across 27 whole-exome sequenced MCL samples. Columns represent individual patients and rows represent recurrently mutated genes. Colors indicate mutation consequence, and the left panel shows the percentage of samples harboring mutations in each gene.

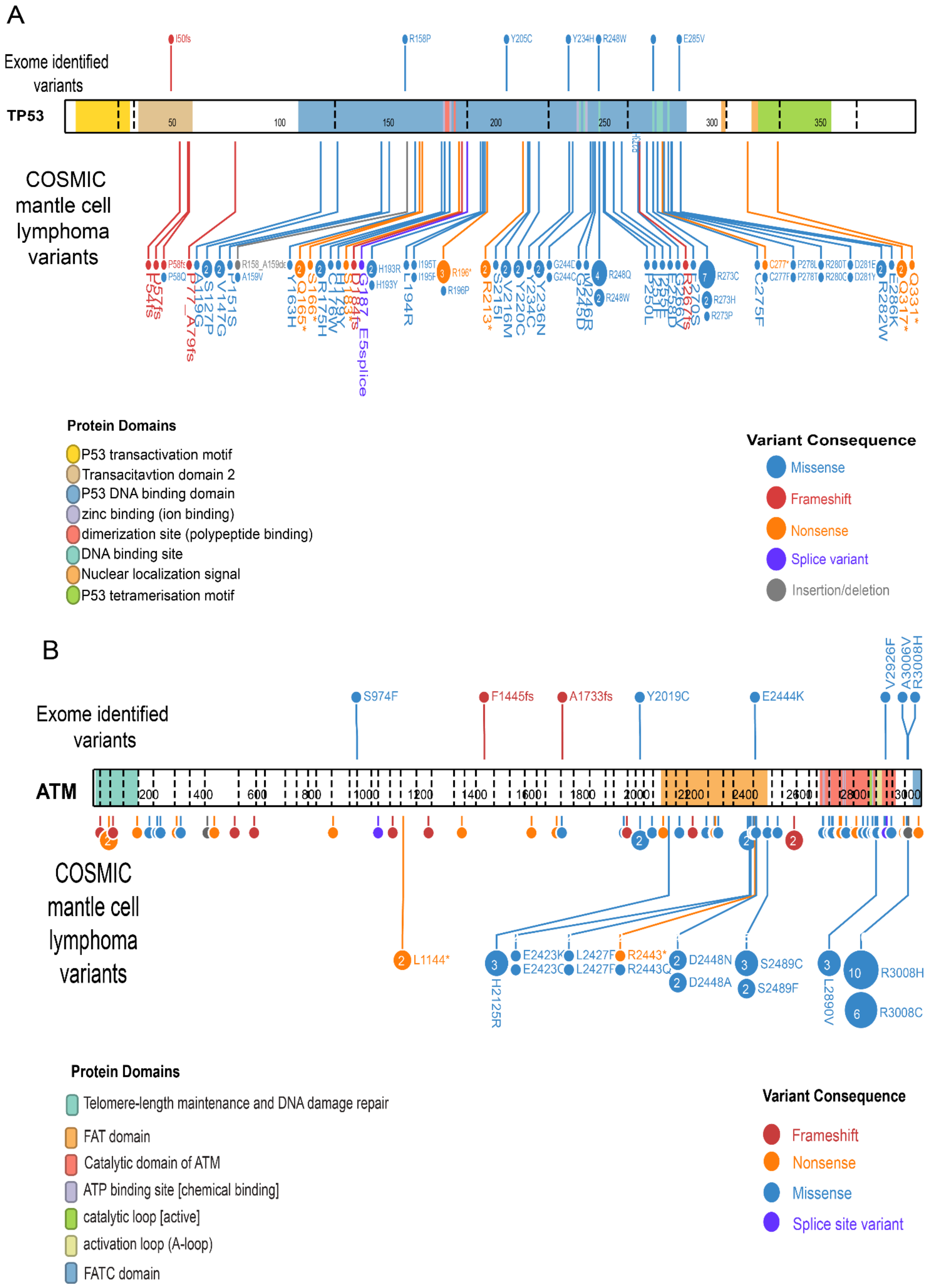
A
Exome identified variants
TP53
COSMIC mantle cell lymphoma variants
Protein Domains
P53 transactivation motif
Transacitavtion domain 2
P53 DNA binding domain
zinc binding (ion binding)
dimerization site (polypeptide binding)
DNA binding site
Nuclear localization signal
P53 tetramerisation motif
Variant Consequence
Missense
Frameshift
Nonsense
Splice variant
Insertion/deletion
B
Exome identified variants
S974F
F1445fs
A1733fs
Y2019C
E2444K
V2926F
A3006V
R3008H
ATM
COSMIC mantle cell lymphoma variants
L1144*
H2125R
E2423K
E2423C
L2427F
L2427F
R2443*
R2443Q
D2448N
D2448A
S2489C
S2489F
L2890V
R3008H
R3008C
Protein Domains
Telomere-length maintenance and DNA damage repair
FAT domain
Catalytic domain of ATM
ATP binding site [chemical binding]
catalytic loop [active]
activation loop (A-loop)
FATC domain
Variant Consequence
Frameshift
Nonsense
Missense
Splice site variant

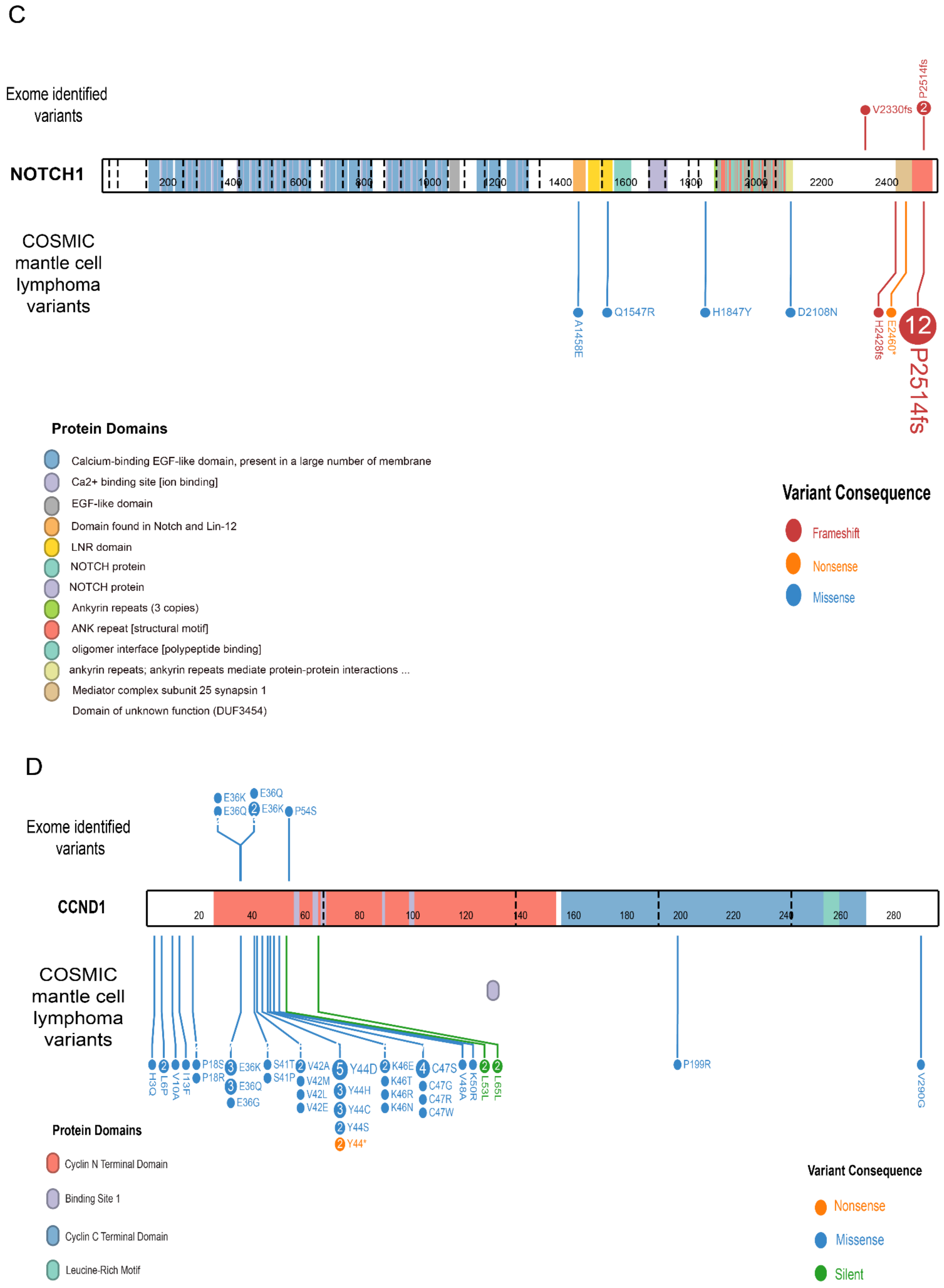
C
Exome identified variants
NOTCH1
V2330fs
P2514fs
COSMIC mantle cell lymphoma variants
A1458E
Q1547R
H1847Y
D2108N
H2428fs
E2460*
P2514fs
Protein Domains
Calcium-binding EGF-like domain, present in a large number of membrane
Ca2+ binding site [ion binding]
EGF-like domain
Domain found in Notch and Lin-12
LNR domain
NOTCH protein
NOTCH protein
Ankyrin repeats (3 copies)
ANK repeat [structural motif]
oligomer interface [polypeptide binding]
ankyrin repeats; ankyrin repeats mediate protein-protein interactions ...
Mediator complex subunit 25 synapsin 1
Domain of unknown function (DUF3454)
Variant Consequence
Frameshift
Nonsense
Missense
D
Exome identified variants
E36K
E36Q
E36Q
E36K
P54S
CCND1
COSMIC mantle cell lymphoma variants
H3Q
L6P
V10A
I13F
P18S
P18R
E36K
E36Q
E36G
S41T
S41P
V42A
V42M
V42L
V42E
Y44D
Y44H
Y44C
Y44S
Y44*
K46E
K46T
K46R
K46N
C47S
C47G
C47R
C47W
V48A
K50R
L53L
L65L
P199R
V290G
Protein Domains
Cyclin N Terminal Domain
Binding Site 1
Cyclin C Terminal Domain
Leucine-Rich Motif
Variant Consequence
Nonsense
Missense
Silent

E

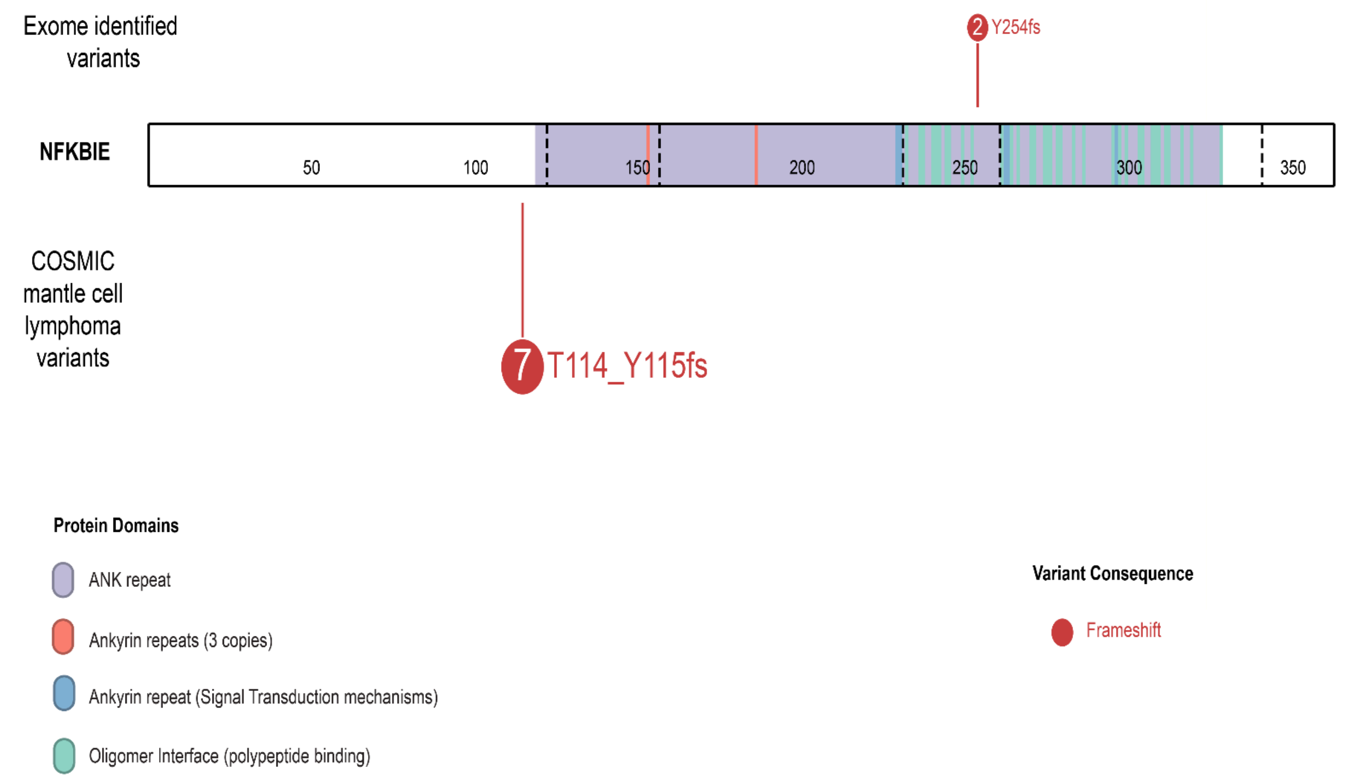

Exome identified variants
2 Y254fs
NFKBIE
50
100
150
200
250
300
350
COSMIC mantle cell lymphoma variants
7 T114_Y115fs
Protein Domains
ANK repeat
Ankyrin repeats (3 copies)
Ankyrin repeat (Signal Transduction mechanisms)
Oligomer Interface (polypeptide binding)
Variant Consequence
Frameshift


F

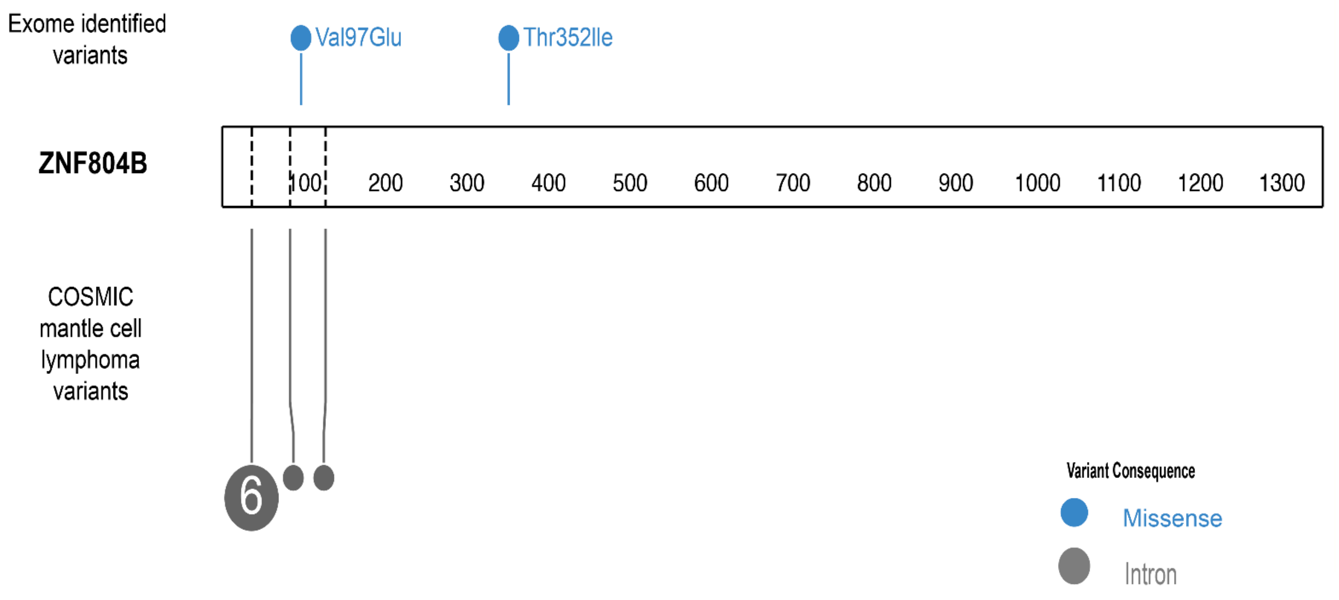

Exome identified variants
Val97Glu
Thr352Ile
ZNF804B
100
200
300
400
500
600
700
800
900
1000
1100
1200
1300
COSMIC mantle cell lymphoma variants
6
Variant Consequence
Missense
Intron

G

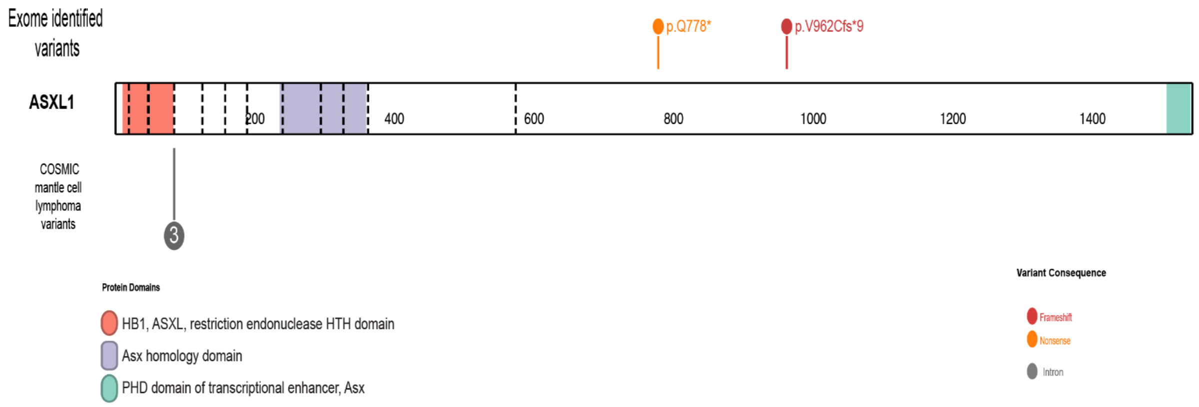


**Supplementary Figure 4. Protein-level distribution of recurrent somatic mutations in MCL.** Lollipop plots showing the positions of coding mutations identified in (A) *TP53*, (B) *ATM*, (C) *NOTCH1*, (D) *CCND1*, (E) *NFKBIE*, (F) *ZNF804B*, and (G) *ASXL1*. Mutations identified in this cohort are shown relative to annotated protein domains and previously reported mantle cell lymphoma variants cataloged in the COSMIC database. The colors indicate variant consequences.

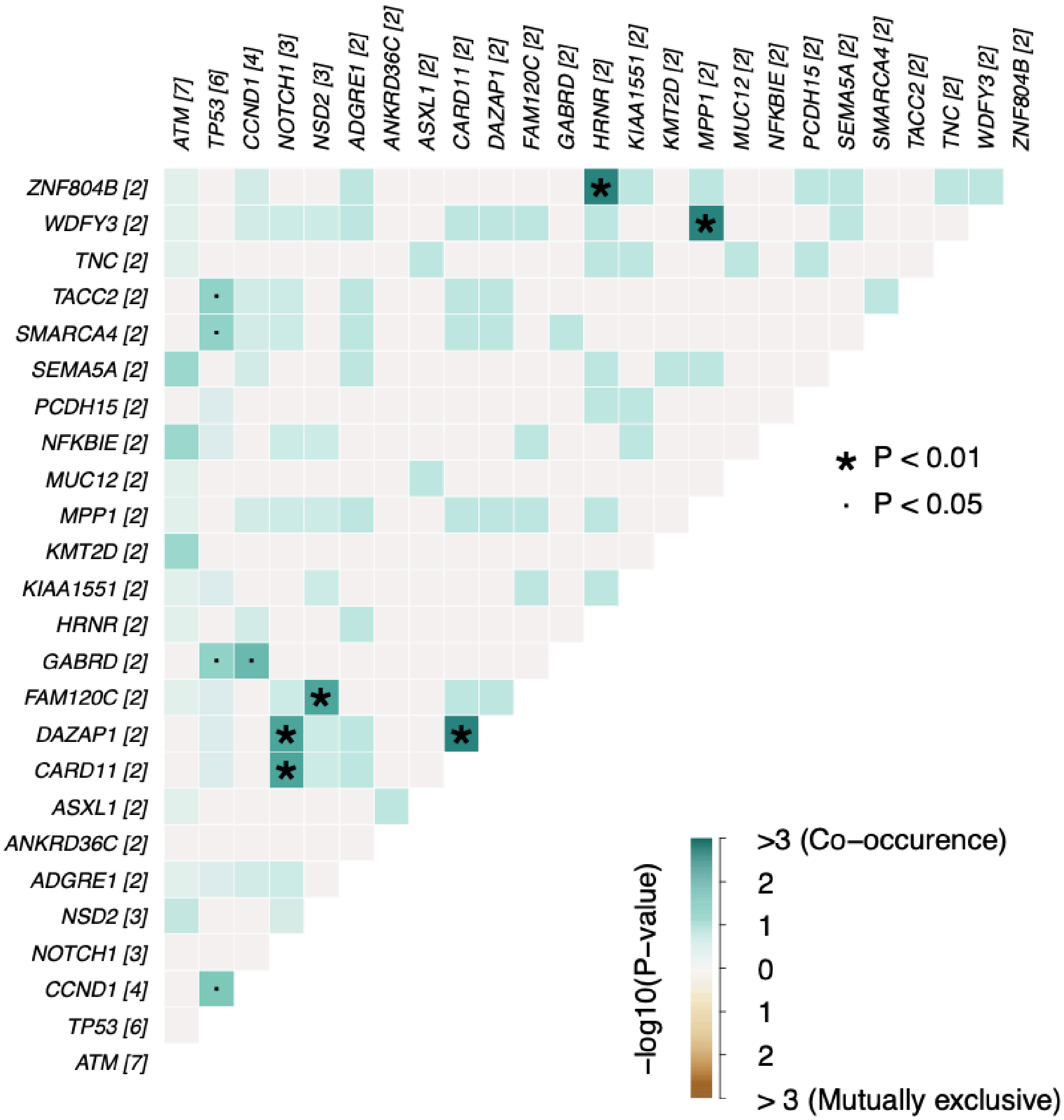


**Supplemental figure 5: Co-occurrence analysis of recurrent somatic SNVs and indels.** Pairwise co-occurrence and mutual exclusivity analysis of recurrently mutated genes identified from SNVs and indels using Fisher's exact test. Cell color represents the strength and direction of association (−log10 P value), with positive values indicating co-occurrence and negative values indicating mutual exclusivity. Symbols denote statistically significant interactions ($P < 0.05$ and $P < 0.01$).

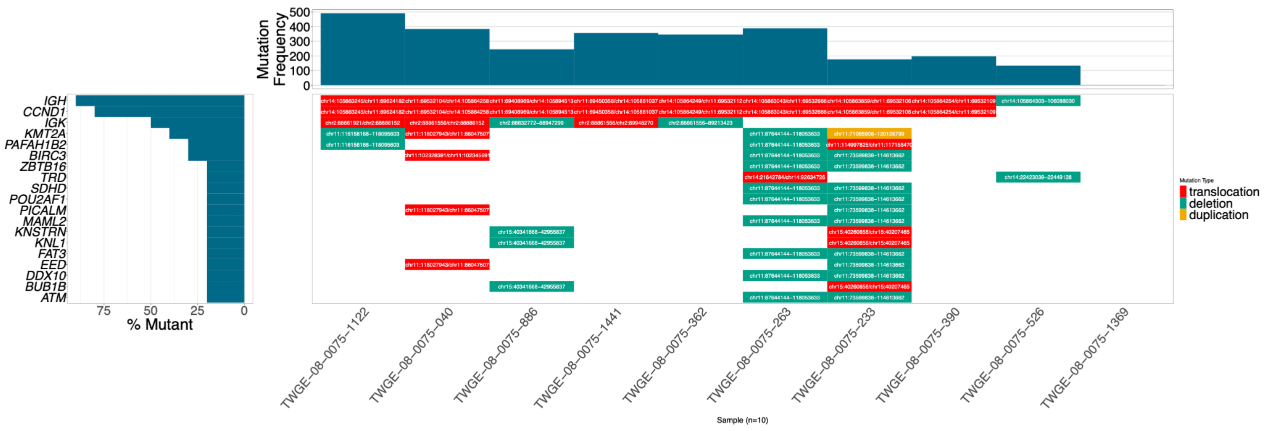


**Supplementary Figure 6. Landscape of recurrent structural variants in the MCL cohort.** Waterfall plot summarizing structural variants identified from 10 WGS MCL samples.Columns represent individual patients and rows represent recurrently affected genes. Colors indicate structural variant class (translocation, deletion, or tandem duplication), while the upper and left panels summarize the number of structural variants per sample and the frequency of alteration for each gene, respectively. The coordinates of each structural variant is indicated for each patient.

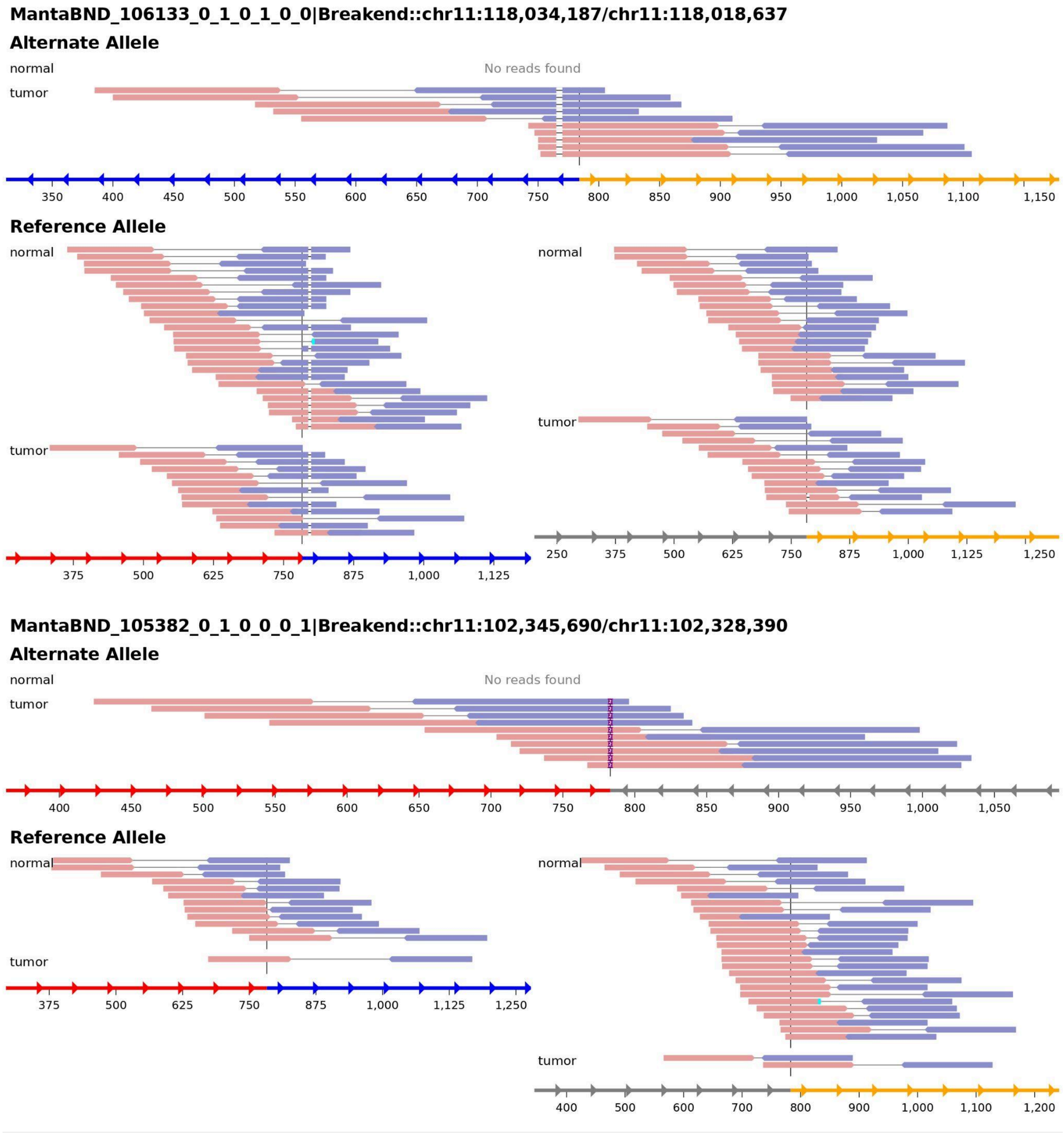


**Supplementary Figure 7. Representative split-read evidence for somatic intrachromosomal inversions involving CCND1.**

Read-level visualization of two intrachromosomal inversion breakpoints identified by whole-genome sequencing. Alternate allele-supporting split reads are detected exclusively in tumor samples and are absent in matched normal controls, supporting the somatic origin of the rearrangements. Reference allele alignments are shown for comparison.
